# Dopamine-induced pruning in Monocyte-Derived-Neuronal-like cells (MDNCs) from patients with Schizophrenia

**DOI:** 10.1101/2021.10.28.21265586

**Authors:** Alfredo Bellon, Vincent Feuillet, Alonso Cortez-Resendiz, Faycal Mouaffak, Lan Kong, L. Elliot Hong, Lilian De Godoy, Therese M. Jay, Anne Hosmalin, Marie-Odile Krebs

## Abstract

The long lapse between the presumptive origin of schizophrenia (SCZ) during early development and its diagnosis in late adolescence has hindered the study of crucial neurodevelopmental processes directly in living patients. Dopamine, a neurotransmitter consistently associated with the pathophysiology of SCZ, participates in several aspects of brain development including pruning of neuronal extensions. Excessive pruning is considered the cause of the most consistent finding in SCZ, namely decreased brain volume. It is therefore possible that patients with SCZ carry an increased susceptibility to dopamine’s pruning effects and that this susceptibility would be more obvious in the early stages of neuronal development when dopamine pruning effects appear to be more prominent. Obtaining developing neurons from living patients is not feasible. Instead, we used Monocyte-Derived-Neuronal-like Cells (MDNCs) as these cells can be generated in only 20 days and deliver reproducible results. In this study, we expanded the number of individuals in whom we tested the reproducibility of MDNCs and deepened the neurostructural comparison between human developing neurons and these neuronal-like cells. Moreover, we studied MDNCs from 12 controls and 13 patients with SCZ. Patients’ cells differentiate more efficiently, extend longer secondary neurites and grow more primary neurites. In addition, MDNCs from a subset of patients expresses less D1R and prune more primary neurites when exposed to dopamine. Haloperidol did not influence our results but the role of other antipsychotics was not examined.

## Introduction

Although schizophrenia (SCZ) is typically diagnosed in late adolescence or early adulthood, this illness is commonly considered as a neurodevelopmental disorder that originates from a combination of genetic predisposition and environmental factors.^1^ This long lapse between SCZ presumptive origin and its diagnosis has hindered the study of early neurodevelopmental processes directly in living patients.

Not surprisingly, the neurodevelopmental process at fault in SCZ remains unidentified. However, evidence collected from adult patients provides some insights. The most consistent finding in SCZ is decreased brain volume^2, 3^ which is attributed to a shrinkage in neuropil.^4–6^ Neuropil reductions are evident across several brain regions.^7–13^ These broad neurostructural abnormalities contradicts the current leading theory suggesting smaller brains in SCZ result from increased pruning of synapses exclusively in one brain region (the prefrontal cortex) and only during the transition between adolescence to adulthood.^4, 6, 14, 15^ Also in conflict with such theory is evidence that the brains of patients are smaller before the clinical onset of SCZ^16–20^ indicating the neuropil is compromised in earlier stages of development. In addition, synapses and spines are not the only affected components of the neuronal structure. Instead, total dendritic length is also reduced^21, 22^ and the number of dendritic branches are likewise decreased.^23, 24^ Consistent with widespread deficits in the neuronal structure, is the variety of neuronal types affected in SCZ, namely; pyramidal,^21–24^ GABAergic,^25^ dopaminergic^12^ and Purkinje cells.^11^ Further support of early and broad neurostructural deficits is that developing neurons generated from patients’ stem cells not yet committed into any particular neuronal type present shorter neuronal extensions.^26, 27^ These widespread defects involving several components of the neuronal structure found in postmortem studies as well as in stem-cells-derived-developing-neurons, suggest that patients with SCZ may carry an increased susceptibility to pruning of neuronal extensions from early stages of neuronal development.

A second consistent pathophysiological aspect in SCZ, but rarely approached from a neurodevelopmental perspective, is its association with dopamine. This relationship relies on several lines of evidence. For instance, at high levels, dopamine produces psychotic symptoms in healthy individuals^28^ and exacerbates psychosis in patients with SCZ.^29, 30^ Accordingly, clinical efficacy of most antipsychotic medications is linked to blocking dopamine receptors.^31, 32^ Neuroimaging studies also support the involvement of dopamine in SCZ. Positron emission tomography (PET) and single-photon emission computed tomography (SPECT) have consistently revealed increased presynaptic dopamine synthesis.^33^ Even individuals at ultra-high risk for psychosis have shown elevated dopamine synthesis capacity.^34^ Dopamine receptors 1 and 2 have also been associated with this illness. The first study to quantify dopamine D_1_ receptors (D1R) in living, unmedicated individuals with SCZ found it decreased.^35^ Follow up results however, have been mixed.^36, 37^ In contrast, reductions in D_2_ receptors (D2R) are commonly encountered.^38^ Overall, the mechanisms of dopamine involvement in the pathophysiology of SCZ remains unsolved. A scarcely explored research avenue in SCZ, is the role of dopamine during early neurodevelopment. Dopamine participates in several aspects of brain development including sculpting the neuronal shape.^39^ *In vivo* studies with animal models of neurodevelopment indicate that activation of D1R decreases neurite extensions.^40–42^ In line with these results are cell culture experiments using developing neurons and neuronal cell lines in which dopamine elicits retraction of neuronal processes, mostly through activation of D1R.^43–46^

Evidence of shorter neuronal extensions in patients with SCZ, together with the regularity in which dopamine has been associated with this psychotic disorder, and the role of dopamine in retracting neuronal extensions, compelled us to propose the following hypothesis: patients with SCZ carry an increased susceptibility to dopamine’s pruning effects. This susceptibility would be more obvious in the early stages of neuronal development when dopamine pruning effects appear to be more prominent. The main challenge to test this hypothesis is that obtaining developing neurons from living patients is not feasible. To circumvent this problem, we used Monocyte-Derived-Neuronal-like Cells (MDNCs).

We have recently developed a protocol to transdifferentiate human circulating monocytes into neuronal-like cells without the need for reprograming.^47^ These MDNCs structurally resemble developing human neurons, conduct electrical activity and express several genes and proteins associated with the inherited predisposition to SCZ. In addition, MDNCs express D1R and retract its neuronal extensions when exposed to dopamine.^47^ Moreover, MDNCs offer two advantages over other cellular models. First, the entire transdifferentiation process from somatic cell to neuronal-like cell takes only 20 days which contrasts with other models that take months to develop.^47^ Second, we have previously shown that MDNCs deliver reproducible results with serial samples from the same individual.^47^ Concerns have been raised about the reproducibility of results with other *in vitro* models.^48, 49^

In this manuscript we expanded the number of individuals and serial samples in which we tested the reproducibility of MDNCs’ results. We then compared if the neurostructure of MDNCs behaves similarly to that of human developing neurons *in vitro*. The neurostructure of MDNCs from 12 controls and 13 patients with schizophrenia was thoroughly studied. Dopamine pruning effects on MDNCs were assessed, as well as the expression of D1R. The potential influence of haloperidol on our results was also studied.

## Materials and Methods

### Subjects

All participants, after receiving full description of the study, gave their informed and written consent. All study procedures were approved by local ethics committees and were in accordance with the Helsinki declaration. Experiments pertaining to the cohort of patients and controls were approved by the ethics committee Ile de France II while experiments on the characterization of MDNCs involving only control individuals were approved by the Institutional Review Board at Penn State University (Study #00006911).

Patients were diagnosed using criteria described in the Diagnostic and Statistical Manual of Mental Disorders, 4^th^ Edition (DSM-IV). Diagnosis was based on clinical interviews and clinical records. Patients were recruited from the Department of Psychiatry at Sainte-Anne Hospital in Paris, France. Healthy controls were recruited through local advertisements. Controls were screened to rule out any past or present history of DSM-IV axis 1 disorders. Only individuals older than 18 years old were recruited.

Thirty-five subjects, 16 controls and 19 patients with SCZ, were recruited for this study. In the SCZ group one patient was diagnosed with schizoaffective disorder and another with pervasive developmental disorder while one control had hemochromatosis (Table 1). One patient and one control provided blood samples in two separate occasions and were included as different subjects in the analysis. Eight individuals, five patients and 3 controls, were excluded from the study due to a mistake in the concentration of growth factors used during the transdifferentiation process. Of the 13 patients included in the analysis, two were not receiving any medications (Table 1). Some results from healthy subjects were included in a prior publication.^47^

**Table 1.** Demographics and medication intake of individuals included in the analysis.

| CONTROLS <n = 12, one tested twice> |  |  |  | SCHIZOPHRENIA <n = 13, one tested twice> |  |  |  |
| --- | --- | --- | --- | --- | --- | --- | --- |
| Diagnosis | Age (range) | Sex | Medications (doses per day) | Diagnosis | Age (range) | Sex | Medications (doses per day) |
| N/A | 18-21 | M | None | Pervasive Developmental Disorder | 18-21 | M | Risperidone 6mg, Valproic Acid 150mg; Methylphenidate 20mg |
| N/A | 26-29 | M | None | Schizophrenia Disorganized type | 18-21 | M* | Haloperidol decanote 150mg/month, Zopiclone 7.5mg, Lithium 800mg, Chlorpromazine 25mg, Cyamemazine 50mg |
| N/A | 26-29 | M* | None | Schizophrenia Undifferentiated type | 26-29 | M | Clozapine 300mg, Aripiprazole 15mg |
| N/A | 26-29 | F | None | Schizophrenia Undifferentiated type | 26-29 | F | Aripiprazole 15mg |
| N/A | 30-33 | M | None | Schizophrenia Undifferentiated type | 26-29 | M | Untreated |
| N/A | 30-33 | M | None | Schizophrenia Undifferentiated type | 30-33 | M | Clozapine 25mg, Duloxetine 120mg |
| N/A | 30-33 | F | None | Schizophrenia Paranoid type | 42-45 | F | Risperidone 6mg, Chlorpromazine 100mg, Oxazepam 150mg, Amimemazine 20mg |
| N/A | 30-33 | M | None | Schizophrenia Disorganized type | 34-37 | M | Valproic Acid ER 1000mg, Aripiprazole 15mg, escitalopram 10mg |
| N/A | 34-37 | M | Loratadine 10mg | Schizophrenia Paranoid type | 34-37 | M | Untreated |
| Hemochromatosis | 46-49 | M | None | Schizoaffective disorder | 42-45 | M | Clozapine 600mg, Valproic Acid 1500mg, Chlorpromazine 300mg, Hydroxyzine 300mg, Diazepam 15mg |
| N/A | 62-65 | M | Clopidogrel 75mg, Previscan 20mg, Rampril 2.5mg, Simvastatin 40mg, Omeprazol 20mg | Schizophrenia Disorganized type | 66-69 | M | Amisulpride 100mg |
| N/A | 26-29 | F | None | Schizophrenia Undifferentiated type | 30-33 | M | Aripiprazole 15mg |
|  |  |  |  | Schizophrenia Paranoid type | 42-45 | M | Haloperidol 30mg, Zopiclone 7.5mg, Chlorpromazine 600mg, Diazepam 30mg |

\*these individuals were tested twice.

### Cell culture

We followed our transdifferentiation protocol described in detail elsewhere.^47^ Briefly, blood samples were processed within 24 hours and when possible shortly after sampling. To calculate the yield, monocyte and peripheral blood mononuclear cells (PBMC) counts were adjusted to 40 ml of blood as for some individuals we obtained only 30ml. PBMCs were isolated from fresh blood using centrifugation over Ficoll-Paque (GE Healthcare, 17-1440-03). A fraction of PBMCs was cultured on fibronectin-coated 25 cm^2^ flasks (13.5 million PBMCs per flask), to generate “PBMC-condition media”. The remaining PBMCs were used for isolation of CD14+ cells (monocytes) by positive immunomagnetic selection (CD14 human Microbeads, Miltenyi Biotec, 130-050-201). CD14+ cells were cultured on fibronectin-coated wells at a concentration of 180,000 cells per cm^2^. Plastic plates and flasks came from BD Falcon (351146, 353043 and 353109). Human fibronectin from plasma (Sigma-Aldrich, F2006) was used at a concentration of 20µg/ml in PBS for coating plates and flasks overnight at 4°C. Macrophage colony-stimulating factor (MCSF) from AbCys (300-25) was added to monocytes right before culturing at a final concentration of 50 ng/ml. All cells were maintained in “Supplemented-DMEM” Dulbecco’s Modified Eagle Medium (DMEM), High Glucose, GlutaMAX (GIBCO, 61965059) in which we added 100 U/mL penicillin; 100 mg/mL streptomycin, 1% nonessential amino acids, 1 mM sodium pyruvate, 10 mM HEPES buffer, (all from Life Technologies) and supplemented with 10% fetal bovine serum (FBS) from GIBCO Performance Plus.

After 4 days in culture at 37°C with 5% CO_2_ pressure, cell media was changed. Fresh Supplemented-DMEM as well as PBMC-conditioned media at a rate of 2:1 were used to replace old media. On day 7, cultured media was again replaced at a ratio of 1:1 (PBMC-conditioned media to supplemented-DMEM). Butylated hydroxyanisole (BHA) (Sigma-Aldrich, B1253) was added to a final concentration of 50 nM. On day 10 media replacement resembled day 7 except for the addition of retinoic acid (RA) (Sigma-Aldrich, R2625) at a final concentration of 16 µM. On day 13, media replacement involved a 1:1 PBMC-condition media to Supplemented-DMEM ratio and adding final concentrations of BHA 50 µM, RA 16 µM, Insulin Growth Factor-1 12.5 ng/ml (Peprotech, 100-11) and Neurotrophin-3 30 ng/ml (Peprotech, 450-03-100). On day 17, cell culture media was not replaced, instead 25 mM Potassium chloride (KCL) was added (Sigma-Aldrich, P5405).

Pictures of cells were taken using a Nikon Eclipse Ti-S/L 100 inverted microscope equipped with a CoolSNAP Myo, 20 MHz, 2.8 Megapixel, 4.54 x 4.54 µm pixels camera and with a Nikon CFI Super fluor 20X DIC prism objective. Pictures were taken immediately after monocyte extraction and then at days 4, 7, 10 and 13 to establish a structural path to differentiation. Pictures were also taken around day 20 (days 19 to 22) when transdifferentiation is completed. These pictures were used to establish differentiation rates. For one of the individuals in whom we collected two serial samples, we observed lower cell concentration with the second sample. When measuring differentiation rates for this subject, we adjusted for low concentration by using the lowest concentration value on the first sample as a threshold. Any pictures from the second sample with a cell concentration lower than the threshold were not included in the differentiation rate analysis for this particular individual.

Treatments with colchicine (Sigma-Aldrich, C9754) and dopamine (Sigma-Aldrich, H8502-259), involved three concentrations for colchicine (0.4µM, 0.5µM and 0.75µM) and two for dopamine (4mM and 5mM). Dopamine was diluted in 1 mg/ml ascorbic acid (Sigma-Aldrich, A4544) dissolved in PBS. To determine the role of D1R in pruning, preincubations with the D1-like receptor antagonist SCH-23390 (Tocris Bioscience, 0925) were conducted for 20 minutes at 5µM. At least 12 different fields in each well were identified via a micro-ruled coverslip (Cellattice CLS5-25D, Nexcelom Bioscience) and pictures were taken before treatments. Cells were then treated with either one of the concentrations for colchicine or dopamine or with medium alone as control. After incubating for one hour at 37°C and 5% CO_2_, pictures of the exact same fields located via the micro-ruled coverslip were taken again. Only neuronal-like cells with at least one primary neurite longer than two times the soma size before treatment were traced. Only the longest primary and the longest secondary neurite for each cell traced were included in the analysis. Cells were traced manually using a semi-automated software called FIJI which is a plugin for Image J an open source image processing program. This same software was used for all structural analyses. Tracing and cell characterization was done blinded.

Human neurons were obtained from ScienCell Research laboratories (1520-10) and cultured following the manufacturer’s instructions. We used ScienCell neuronal medium (1521) and plastic plates were coated with Poly-L-Lysine, 1 mg/ml (0403). Pictures of human neurons were taken after 5 days in culture. Human neurons were traced as described for neuronal-like cells. Only neurons with at least one primary neurite longer than two times the soma size were traced. Only the longest primary and the longest secondary neurite for each cell traced were included in the analysis. Neurons in which the exact ending of neurites was uncertain were not traced.

### Flow cytometry

Cells were detached by incubating them at 37°C with Trypsin-EDTA (0.05%), phenol red (ThermoFisher Scientific, 25300). Incubation times were 4 minutes for neuronal-like cells and 15 minutes for macrophages as previously reported.^47, 50, 51^ Macrophages were obtained from PBMCs cultured to generate PBMC-condition media. After washing them, cells were treated with Blue Live/Dead Stain Kit (Life Technology, L23105) or 4’,6-Diamidino-2-Phenylindole, Dihydrochloride (DAPI) (ThermoFisher Scientific, D1306) to exclude dead cells from analysis. Saturation was performed with either human group AB serum or Human BD Fc block (BD Pharmingen, 564220). The following antibodies were incubated for 20 minutes on ice for extracellular labeling: mouse IgG2a anti-human CD14-Qdot-655 (1/20, Invitrogen, Q10056) and mouse IgG2b anti-human Dopamine Receptor 1-PE (1/20, BioLegend, 366404). Cells were then fixed with paraformaldehyde (PFA) 4%. For intracellular proteins, permeabilization was done with Triton X-100 0.2%. Rabbit anti-Nestin (1/200, Millipore, AB5922) was incubated 45 minutes on ice. Alexa Fluor 488 Goat anti-Rabbit (1/100, ThermoFisher Scientfic, A-11070) was used as secondary antibody and incubated 20 minutes on ice. Events were acquired using a FACS CANTO, Fortessa or a BD LSR II flow cytometer and analyzed using Diva (Version 6.1.1, BD Biosciences) followed by FlowJo (Version 10.1r7; TreeStar).

### Statistical Analysis

We conducted a descriptive analysis to examine the distributions of measures from MDNCs for healthy controls and SCZ patients. Subject-level measurements for MDNCs were obtained by first averaging the picture-level data for each sample and then averaging the sample-level data within each subject. Differentiation efficiency and structural parameters were summarized at the subject-level and compared between study groups using analysis of variance (ANOVA) or two-sample t-tests. The data at picture- or sample-level were also analyzed with a mixed model analysis to account for correlations of repeated measures within subjects. To compare pruning effects of experimental conditions between study groups, we adjusted for baseline retraction (response under control conditions), differentiation efficiency, and structure at baseline because these factors may affect the pruning effects of each treatment condition (dopamine & colchicine). Since different samples were used to measure these factors, we were unable to perform analysis with picture level data. Therefore, we ran linear regression analysis based on subject-level averaged data adjusting for these factors in the models as covariates. Least square means and standard errors were estimated from the models for each study group.

Non-parametric Mann-Whitney test was used to make pairwise comparisons between MDNCs and HDNs as well as for determining expression levels of D1R and CD14. The Kruskal-Wallis test was used to make comparisons between MDNCs incubated under control conditions, vehicle and haloperidol. P values lower or equal to 0.05 were considered significant.

## Results

### Reproducibility of results with MDNCs

We have previously shown that the neuronal structure of MDNCs transdifferentiated from two serial blood samples from four healthy men were not statistically different.^47^ Here we expand our cohort to eight healthy individuals, five men and three women (Table 2). We also increased the number of samples in two subjects from two to three serial samples (Table 2). For each parameter studied, we compared the distributions between samples within each subject using Kolmogorov-Smirnov Tests and construct 95% confidence intervals for sample means and mean differences between samples (Fig. 1A).

**Figure 1.**
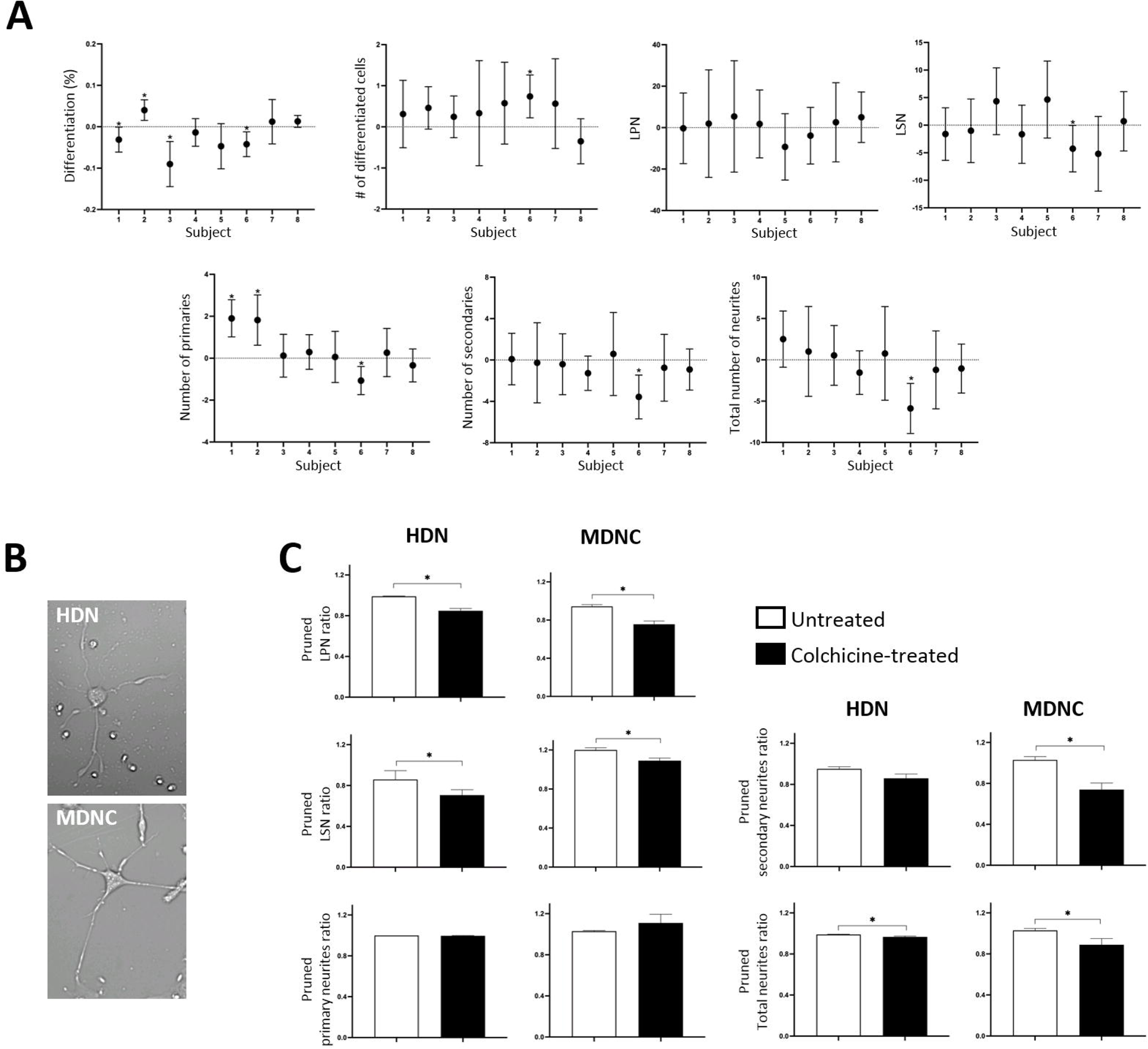
Reproducibility of results with MDNCs and structural comparison with human developing neurons. **(A)** Confidence interval plots in which dots represent means of differences between samples and error bars correspond to 95% confidence interval constructed using the Kolmogorov-Smirnov Test. The parameters presented are: differentiation percentage (subject 1, sample 1 (S1) *n* = 336 (cells), S2 *n* = 433; subject 2, S1 *n* = 284, S2 *n* = 523; subject 3, S1 *n* = 167, S2 *n* = 133, S3 *n* = 464; subject 4, S1 *n* = 556, S2 *n* = 434, S3 *n* = 255; subject 5, S1 *n* = 251, S2 *n* = 193; subject 6, S1 *n* = 797, S2 *n* = 413; subject 7, S1 *n* = 167, S2 *n* = 180; subject 8, S1 *n* = 1027, S2 *n* = 1531), number of differentiated cells (subject 1, S1 *n* = 27, S2 *n* = 45; subject 2, S1 *n* = 20, S2 *n* = 19; subject 3, S1 *n* = 16, S2 *n* = 23, S3 *n* = 38; subject 4, S1 *n* = 36, S2 *n* = 33, S3 *n* = 30; subject 5, S1 *n* = 25, S2 *n* = 25; subject 6, S1 *n* = 74, S2 *n* = 54; subject 7, S1 *n* = 28, S2 *n* = 28; subject 8, S1 *n* = 44, S2 *n* = 57), longest primary neurite (LPN) (subject 1, S1 *n* = 27, S2 *n* = 45; subject 2, S1 *n* = 20, S2 *n* = 19; subject 3, S1 *n* = 16, S2 *n* = 23, S3 *n* = 38; subject 4, S1 *n* = 36, S2 *n* = 33, S3 *n* = 30; subject 5, S1 *n* = 25, S2 *n* = 39; subject 6, S1 *n* = 74, S2 *n* = 54; subject 7, S1 *n* = 28, S2 *n* = 28; subject 8, S1 *n* = 44, S2 *n* = 57), longest secondary neurite (LSN) (subject 1, S1 *n* = 27, S2 *n* = 45; subject 2, S1 *n* = 20, S2 *n* = 19; subject 3, S1 *n* = 16, S2 *n* = 23, S3 *n* = 38; subject 4, S1 *n* = 35, S2 *n* = 33, S3 *n* = 30; subject 5, S1 *n* = 25, S2 *n* = 39; subject 6, S1 *n* = 67, S2 *n* = 54; subject 7, S1 *n* = 28, S2 *n* = 28; subject 8, S1 *n* = 42, S2 *n* = 56), number of primary neurites (subject 1, S1 *n* = 26, S2 *n* = 37; subject 2, S1 *n* = 12, S2 *n* = 13; subject 3, S1 *n* = 14, S2 *n* = 19, S3 *n* = 32; subject 4, S1 *n* = 22, S2 *n* = 25, S3 *n* = 23; subject 5, S1 *n* = 25, S2 *n* = 32; subject 6, S1 *n* = 51, S2 *n* = 45; subject 7, S1 *n* = 26, S2 *n* = 23; subject 8, S1 *n* = 38, S2 *n* = 48), number of secondary neurites and number of total neurites (same *n* for both) (subject 1, S1 *n* = 27, S2 *n* = 45; subject 2, S1 *n* = 20, S2 *n* = 19; subject 3, S1 *n* = 16, S2 *n* = 23, S3 *n* = 38; subject 4, S1 *n* = 36, S2 *n* = 33, S3 *n* = 30; subject 5, S1 *n* = 25, S2 *n* = 39; subject 6, S1 *n* = 74, S2 *n* = 54; subject 7, S1 *n* = 28, S2 *n* = 28; subject 8, S1 *n* = 44, S2 *n* = 57). \**P* = or < 0.05. **(B)** Light microscopy photographs of monocyte-derived-neuronal-like cells (MDNC) and human developing neurons (HDN) in culture for 5 days (20x original magnification). Scale bar = 20µm. **(C)** Bar graphs showing the structural response to colchicine in HDNs and MDNCs. Data are expressed as ratios between the number of each measured structural parameter studied (LPN, LSN, number of primary and secondary neurites and total number of neurites) at baseline and after an hour of incubation either under control conditions or after treatment with colchicine 0.5µM. Statistics are given as mean ± SEM. Differences were assessed using the Mann-Whitney test. Experiments for HDN come from 13 wells for each condition obtained from two different vials of human neurons. HDN for LPN, number of primary neurites and total number of neurites, control, *n* = 267 and colchicine, *n* = 261. For LSN control, *n* = 90 and colchicine, *n* =125 and for number of secondary neurites control, *n* = 90 and colchicine, *n* =123. Control MDNCs came from 8 donors and MDNCs treated with colchicine from 4 donors. MDNCs for LPN, number of primary neurites and total number of neurites, *n* = 656 control, *n* = 401 colchicine. MDNCs for LSN, *n* = 571 control, *n* = 267 colchicine. MDNCs for number of secondary neurites, *n* = 611 control, *n* = 357. \**P* = or < 0.05.

**Table 2.**
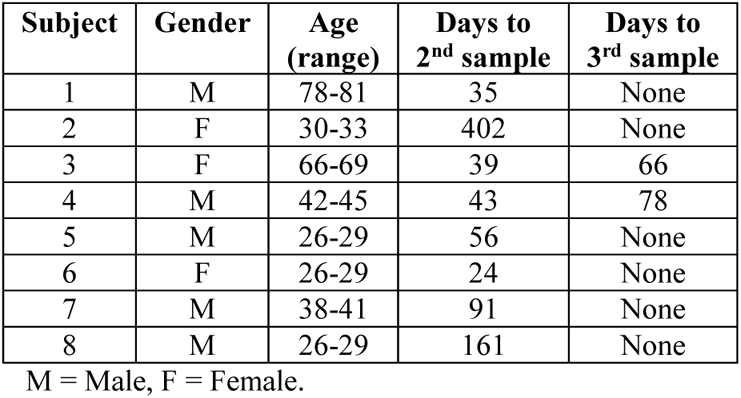
Demographics and number of samples from individuals recruited for experiments on reproducibility.

Differentiation percentage established by cellular phenotype yield significant differences between serial samples for subjects 1 (difference −0.031, 95% confidence interval (CL) −0.061 to −0.0009, *P* = 0.04), 2 (difference 0.040, 95% CL 0.015 to 0.065, *P* = 0.003), 3 (difference −0.090, 95% CL −0.145 to −0.036, *P* = 0.002), and 6 (difference −0.042, 95% CL −0.072 to −0.012, *P* = 0.007) but not for the other healthy individuals, as shown in figure 1A and Supplemental Table 1. The number of differentiated cells evidence statistical significant differences between samples only for subject 6 (difference 0.741, 95% CL 0.220 to 1.262, *P* = 0.006). No differences were observed in all other individuals (Fig. 1A & Supplemental Table 2).

Different neurostructural parameters of MDNCs were assessed: longest primary neurite (LPN), longest secondary neurite (LSN), number of primary neurites, number of secondary neurites, and number of all neurites (including primary, secondary, tertiary and quaternary). There are no statistical differences in LPN in any of the 8 healthy subjects (Fig. 1A & Supplemental Table 3). For LSN, only subject 6 exhibits statistical changes between samples (difference −4.26, 95% CL −8.48 to −0.054, *P* = 0.04) whereas all other participants do not (Fig. 1A & Supplemental Table 4). Number of primary neurites reveals differences in subjects 1 (difference 1.9, 95% CL 1.02 to 2.79, *P* = 0.0001), 2 (difference 1.82, 95% CL 0.62 to 3.02, *P* = 0.005) and 6 (difference −1.07, 95% CL −1.74 to 0.4, *P* = 0.002) but not in subjects 3, 4, 5, 7 & 8 (Fig. 1A & Supplemental Table 5). Number of secondary neurites reveals differences only for subject 6 (difference −3.57, 95% CL −5.69 to 1.46, *P* = 0.001) whereas no statistical changes are evident in the rest of the cohort (Fig. 1A & Supplemental Table 6). Statistical differences for total number of neurites were limited to subject 6 (difference −5.88, 95% CL −8.91 to −2.85, *P* = 0.0002). All serial samples from the other 7 participants evidence no statistical change (Fig. 1A & Supplemental Table 7). Therefore, these data indicates that MDNCs deliver reproducible results for number of differentiated cells, LPN, LSN, number of secondary neurites and total number of neurites. Number of primary neurites delivers reproducible results in 5 out of 8 individuals while percentage of differentiated cells in 4 out of 8 subjects (Fig. 1A).

### Human Developing Neurons versus MDNCs

In a previous publication we showed that the structure of MDNCs is comparable to the structure of Human Developing Neurons (HDNs) after 5 days in culture^47^ (Fig. 1B). Here we compare how HDNs and MDNCs respond to colchicine 0.5µM. Colchicine was chosen because it is well-known within the neuroscientific field for its ability to elicit pruning of neuronal extensions.^52^ In addition, the mechanism of action of colchicine is well-understood and it is independent of membrane receptors as it acts by directly depolymerizing microtubules.^53^

Under culture control conditions both HDNs and MDNCs retract their LPN slightly after one hour of incubation in medium alone (Fig. 1C). Treatment with colchicine 0.5µM for one hour leads to additional retraction of LPN that is comparable in both cell types. HDNs present a net LPN retraction of 14%, which is significantly different when compared to the level of retraction under control conditions (0.99 ± 0.004 for control, and 0.848 ± 0.023 for colchicine, *P* < 0.00001). Likewise, MDNCs present an LPN net retraction of 19% when incubated with colchicine (0.943 ± 0.019 for control, and 0.756 ± 0.035 for colchicine, *P* = 0.0067). The behavior of secondary neurites from HDNs and MDNCs differs. Under control conditions, HDNs retract their LSN by 15% while MDNCs’ LSN grow by 20% (Fig.1C). Exposure to colchicine 0.5µM elicits even further retraction of HDNs’ LSN by another 15% (0.859 ± 0.086 for control, and 0.706 ± 0.053 for colchicine, *P* = 0.05) whereas for MCNCs, colchicine prevents growth of LSN by 11% (1.2 ± 0.022 for control, and 1.09 ± 0.026 for colchicine, *P* = 0.019).

Pruning of primary neurites is comparable between HDNs and MDNCs. Neither HDNs nor MDNCs prune any of its primary neurites after one hour of culture under control conditions (Fig. 1C). Treatment with colchicine leads to no statistical significant change in the number of primary neurites for either HDNs or MDNCs (HDNs, 1 ± 0.00 for control, and 0.997 ± 0.003 for colchicine, *P* = 0.75; MDNCs, 1.03 ± 0.006 for control, and 1.11 ± 0.084 for colchicine, *P* = 0.88). Pruning of secondary neurites is different between HDNs and MDNCs (Fig. 1C). Control conditions cause minimal pruning of secondary neurites in HDNs, while MDNCs do not lose secondary neurites. Incubation with colchicine eliminates an additional 10% in the number of secondary neurites for HDNs (0.951 ± 0.021 for control, and 0.859 ± 0.042 for colchicine, *P* = 0.12) but this difference is not statistically significant. For MDNCs, colchicine elicits a 25% reduction in the number of secondary neurites and this effect reaches statistical significance (1.03 ± 0.032 for control, and 0.74 ± 0.064 for colchicine, *P* = 0.008). When all neurites are included in the analysis (primary, secondary, tertiary and quaternary neurites), HDNs and MDNCs present comparable results (Fig. 1C). No changes are seen under control conditions for both cell types. Treatment with colchicine however, removes a small but statistically significant number of total neurites for HDNs (0.99 ± 0.004 for control, and 0.967 ± 0.008 for colchicine, *P* = 0.04) and slightly higher number of pruned total neurites for MDNCs (1.03 ± 0.019 for control, and 0.89 ± 0.06 for colchicine, *P* = 0.03). Altogether, these results show that, in addition to having comparable structure, HDNs and MDNCs behave similarly following colchicine treatment in terms of pruning of neurites which makes MDNCs relevant for studying at least some neurostructural aspects of early neuronal development.

### Monocytes from controls versus SCZ at baseline and during early stages of transdifferentiation

Twenty-five individuals, 12 controls (CTL) and 13 patients with SCZ were included in the analysis. Two patients with SCZ were not taking any medications (Table 1). The other 11 patients were receiving antipsychotics as well as other psychotropic medications (Table 1). Two-sample t-tests assuming unequal variances were used to compare mean differences between groups on continuous variables and Fisher’s exact tests were used for categorical variables. No statistical differences between groups are evidenced in gender (CTL, 77% men; SCZ, 86% men; *P* = 0.64), age (CTL, 32.6 ± 3.2; SCZ, 33.2 ± 3.4; *P* = 0.88), total number of monocytes per blood sample (CTL, 6.58 ± 0.72 million; SCZ, 8.02 ± 1.17 million; *P* = 0.30) or, total number of peripheral blood mononuclear cells (PBMCs) per blood sample (CTL, 71.65 ± 6.06 million; SCZ, 59.28 ± 3.76 million; *P* = 0.09) (Table 3). The percentage of monocytes within PBMCs presents a small but statically significant difference between groups (CTL, 9.6 ± 0.64%; SCZ, 12.8 ± 1.3%; *P* = 0.04) (Table 3). For one control we did not have access to the number of PBMCs or monocytes.

**Table 3.** Monocytes and peripheral blood mononuclear cells at baseline in patients and controls.

|  | **Controls**<br>**(*n* = 13)** | **Schizophrenia**<br>**(*n* = 14)** | ***P* value** |
| --- | --- | --- | --- |
| **Sex** | 77% men | 86% men | *P* = 0.64 |
| **Age**<br>**(mean ± SEM)**<br>**(range)** | 32.6 ± 3.2<br>(19-65 years) | 33.2 ± 3.4<br>(19-67 years) | *P* = 0.88 |
| **Number of**<br>**Monocytes**<br>**(mean ± SEM)**<br>**(range)** | 6.58 ± 0.72<br>(3.8-12.3 million) | 8.02 ± 1.17<br>(2-17 million) | *P* = 0.30 |
| **Number of**<br>**\*PBMCs**<br>**(mean ± SEM)**<br>**(range)** | 71.65 ± 6.06<br>(43-101.5 million) | 59.28 ± 3.76<br>(39-88 million) | *P* = 0.09 |
| **Percentage of**<br>**Monocytes**<br>**(mean ± SEM)**<br>**(range)** | 9.6 ± 0.64%<br>(6.3-13.1%) | 12.8 ± 1.3%<br>(5-21.7%) | *P* = 0.04 |

\*PBMCs = Peripheral blood mononuclear cells.

After isolating monocytes from blood samples, we followed our 20-day transdifferentiation protocol.^47^ As previously described, monocytes undergo four structural stages before reaching transdifferentiation into neuronal-like cells,^47^ including: 1) rounded cells (RC); 2) standard macrophages (SM), 3) fibroblastic shape (FS) and 4) uncharacterized cells (UC), which include cells that do not fit into any of the other 3 descriptions (Fig. 2A). These structural stages were quantified through microphotographs taken from one cell culture well on days when media was changed, namely; days 4, 7, 10 and 13.

**Figure 2.**
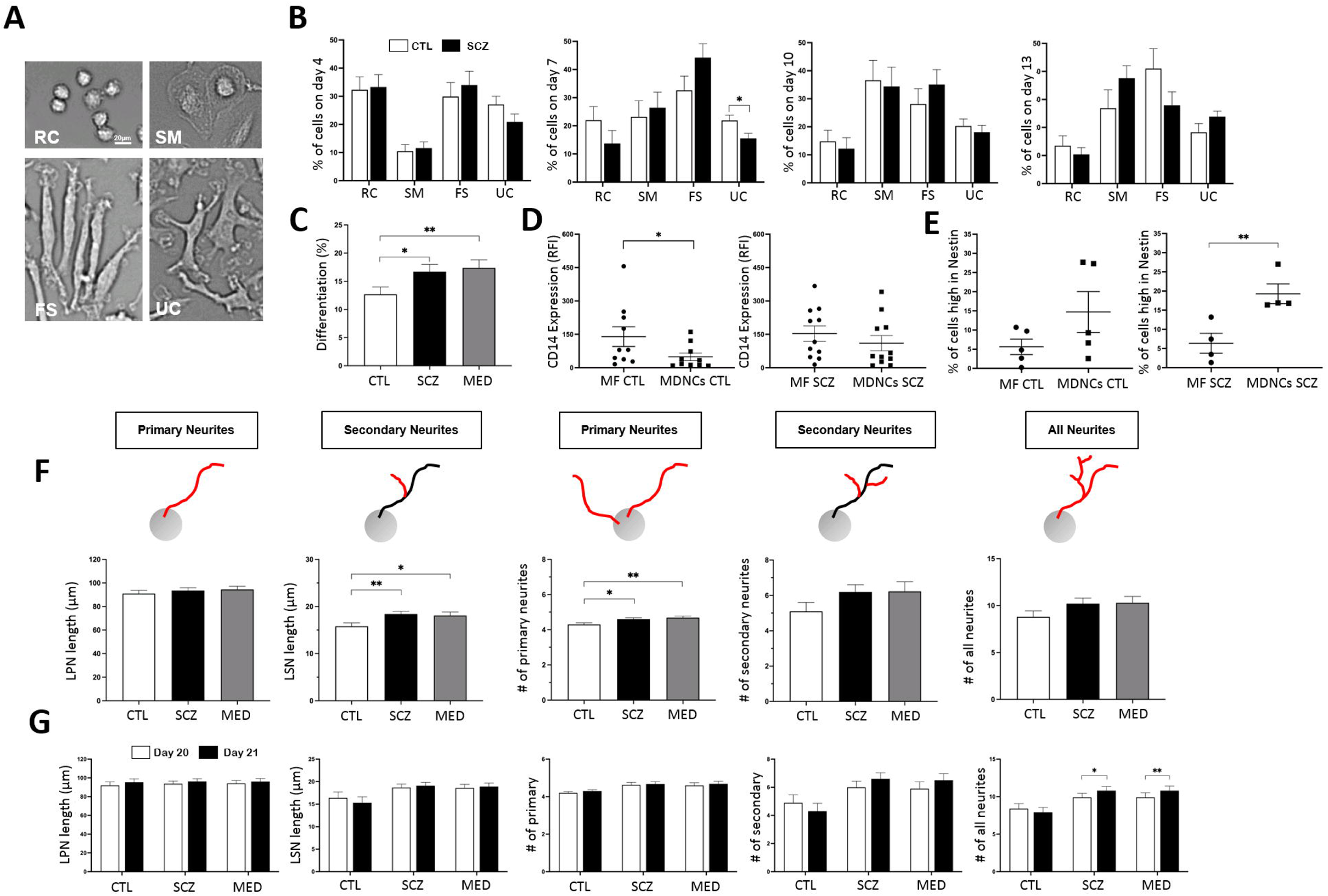
MDNCs from controls (CTL) versus patients with schizophrenia (SCZ). **(A)** Representative light microscopy photographs of the four different morphologies characterized during transdifferentiation: rounded cells (RC), standard macrophages (SM), fibroblastic shape (FS) and uncharacterized (UC) (20x original magnification). Scale bar = 20µm. **(B)** Bar graphs showing variations between controls and patients with schizophrenia in the percentage of each of the four cell morphologies on day 4, 7, 10 and 13 of transdifferentiation. Repeated ANOVAs based on the mixed model framework were performed to examine group differences for each day. Data are given as mean ± SEM. Cells from 13 CTL and 14 SCZ were characterized for day 4, 7 & 10 and 5 CTL and 11 SCZ for day 13. Cells characterized on day 4, CTL *n* = 8437, SCZ *n* = 8204; on day 7, CTL *n* = 7783, SCZ *n* = 7609; on day 10, CTL *n* = 7125, SCZ *n* = 6830; and on day 13, CTL *n* = 2475, SCZ *n* = 5342. \**P* = or < 0.05. **(C)** Bar graphs contrasting the percentage of differentiated cells between CTL, SCZ and only medicated patients (MED). Differentiation was obtained via cell phenotype. To determine differences between groups, multilevel mixed models to account for correlations of repeated measures at subject level and sample level were used, followed by a two-sample t-test. Data are given as mean ± SEM. Cells from 13 CTL and 14 SCZ were included in the analysis. Characterized cells for CTL, *n* = 32791; SCZ *n* = 37281; and for MED *n* = 32986. \**P* = or < 0.05, \*\**P* < 0.03. **(D)** Dot plots contrasting expression of CD14 in macrophages (MF) versus MDNCs from either CTL or SCZ. Differences were assessed using the Mann-Whitney test. Data are given as mean ± SEM. Cells from 10 CTL and 11 SCZ were included in the analysis. \**P* = or < 0.05. **(E)** Dot plots contrasting the percentage of cells expressing high levels of nestin in macrophages (MF) versus MDNCs from either CTL or SCZ. Differences were assessed using the Mann-Whitney test. Data are given as mean ± SEM. Cells from 2 CTL and 4 SCZ were included in the analysis. For one of the CTL four samples were obtained. \**P* = or < 0.05. **(F)** Bar graphs contrasting several structural parameters at baseline between CTL, SCZ and MED. Structural parameters studied were; longest primary neurite (LPN), longest secondary neurite (LSN), number of primary neurites, number of secondary neurites and total number of neurites. To determine differences between groups, multilevel mixed models to account for correlations of repeated measures at subject level and sample level were used, followed by a two-sample t-test. Data are given as mean ± SEM. MDNCs from 13 CTL, 14 SCZ and 12 MED were included in the analysis. MDNCs traced for CTL *n* = 3933; SCZ *n* = 6144; and MED *n* = 5601. \**P* = or < 0.05, \*\**P* < 0.03. **(G)** Bar graphs contrasting several structural parameters on day 20 versus day 21 from CTL, SCZ and MED. Structural parameters studied were; longest primary neurite (LPN), longest secondary neurite (LSN), number of primary neurites, number of secondary neurites and total number of neurites. Two-sample t-tests was used to determine differences between CTL vs. SCZ and CTL vs. MED after a mixed model analysis was performed to account for correlations of repeated measures within subjects. Data are given as mean ± SEM. MDNCs from 7 CTL, 11 SCZ and 10 MED were included in the analysis. MDNCs traced for LSN for CTL, day 20 *n* = 1090 and day 21 *n* = 1091; for all other structural parameters; CTL, day 20 *n* = 1189 and day 21 *n* = 1213; for LSN for SCZ, day 20 *n* = 2062 and day 21 *n* = 2152; for all other structural parameters day 20 *n* = 2193 and day 21 *n* = 2254; for LSN for MED, day 20 *n* = 1985 and day 21 *n* = 2018; for all other structural parameters day 20 *n* = 2114 and day 21 *n* = 2118. \**P* = or < 0.05, \*\**P* < 0.03.

Means and standard deviations were summarized at subject level by study groups. Repeated ANOVAs based on the mixed model framework were performed to examine group differences for each day. Random effect was specified in the models to account for correlations between microphotographs within each subject or variation among subjects. With this approach, the structural path to transdifferentiation between patients and controls shows that there are no statistical differences on days 4, 10 and 13 on any of the structural stages (Fig. 2B & Supplemental Table 8). On day 7, patients evidence less uncharacterized cells while the rest of the structural stages do not differ from controls (Fig. 2B & Supplemental Table 8).

### MDNCs from CTL versus SCZ

After twenty days in culture following our transdifferentiation, around 13% of monocytes acquire a neuronal morphology characterized by the presence of a well-defined soma and long thin neurites^47^ (Fig. 1B). We have previously shown that these monocyte-derived-neuronal-like cells (MDNCs) express a variety of neuronal markers and conduct electrical activity.^47^ In order to compare the percentage of differentiated cells between groups, we collected data through microphotographs taken from multiple samples of each subject. We used multilevel mixed models to account for correlations of repeated measures at subject level and sample level, followed by a two-sample t-test. This approach reveals that the percentage of cells that acquire a neuronal morphology is higher in patients with SCZ than in CTL (Fig. 2C). Such statistical difference is still present when only medicated patients (MED) are included in the analysis (CTL, 12.7 ± 1.3%; SCZ, 16.7 ± 1.3%; *P* = 0.04; MED, 17.4 ± 1.4%; *P* = 0.027) (Fig. 2C).

In a prior publication we showed that MDNCs reduce or even completely abolish their expression of CD14,^47^ a surface marker for monocytes and macrophages. It is therefore expected that MDNCs will express significantly lower CD14 if compared with macrophages from the same individual. MDNCs from CTL behave as predicted (MF, 140 ± 44; MDNCs, 49 ± 17; *P* = 0.03) (Fig. 2D). In contrast, MDNCs from SCZ do not (MF, 154 ± 34.5; MDNCs, 110 ± 33.8; *P* = 0.25) (Fig. 2D). Expression of nestin is also different between groups. The percentage of MDNCs expressing high levels of nestin is higher than in macrophages in both cohorts but for CTL this difference is not statistically significant (CTL, MF, 5.6 ± 2%; MDNCs, 14.7 ± 5.3%; *P* = 0.4; SCZ, MF, 6.3 ± 2.6%; MDNCs, 19.2 ± 2.5%; *P* = 0.01) (Fig. 2E). Likewise, the neurostructure of MDNCs reveals differences. MDNCs from patients with SCZ are structurally more complex than cells from CTL (Fig. 2F). While no differences are evident when comparing LPN between CTL and either SCZ or MED (CTL, 91 ± 2.6µm; SCZ, 93.5 ± 2.4µm; *P* = 0.49, MED, 94.5 ± 2.6µm; *P* = 0.36), MDNCs from patients extend longer secondary neurites (CTL, 15.8 ± 0.7µm; SCZ, 18.4 ± 0.6µm; *P* = 0.02, MED, 18.1 ± 0.7µm; *P* = 0.04) and grow more primary neurites (CTL, 4.3 ± 0.09; SCZ, 4.6 ± 0.08; *P* = 0.04, MED, 4.6 ± 0.09; *P* = 0.01) and the same is true for MED (Fig. 2F). The number of secondary neurites (CTL, 5.1 ± 0.5; SCZ, 6.2 ± 0.4; *P* = 0.17, MED, 6.2 ± 0.5; *P* = 0.19) as well as the number of all neurites, including; primary, secondary, tertiary, quaternary and quinary (CTL, 8.8 ± 0.6; SCZ, 10.2 ± 0.6; *P* = 0.11, MED, 10.3 ± 0.6; *P* = 0.13) does not show statistical differences between groups (Fig. 2F).

Since transdifferentiation occurs after 20 days in culture, the vast majority of MDNCs structural analyses were conducted between days 20 and 21. We analyzed whether there are structural differences between MDNCs at day 20 (D20) versus day 21 (D21) (Fig. 2G). Only those individuals in each group for whom we have microphotographs of MDNCs at D20 and D21 were included in the analysis. MDNCs from controls do not show structural changes in any of the parameters studied namely; LPN (D20, 92 ± 3.6µm; D21, 95.3 ± 3.6µm; *P* = 0.23), LSN (D20, 16.4 ± 1.3µm; D21, 15.3 ± 1.3µm; *P* = 0.25), number of primary neurites (D20, 4.2 ± 0.07; D21, 4.3 ± 0.07; *P* = 0.20), number of secondary neurites (D20, 4.9 ± 0.5; D21, 4.3 ± 0.5; *P* = 0.16), and total number of neurites (D20, 8.4 ± 0.6; D21, 7.9 ± 0.6; *P* = 0.29). In contrast, MDNCs from SCZ evidence a higher number of total neurites on D21 when compared to D20 (D20, 9.9 ± 0.5; D21, 10.8 ± 0.5; *P* = 0.02), while the rest of the structural parameters including; LPN (D20, 93.7 ± 2.9µm; D21, 96.2 ± 2.9µm; *P* = 0.08), LSN (D20, 18.7 ± 0.75µm; D21, 19.1 ± 0.74µm; *P* = 0.60), number of primary neurites (D20, 4.63 ± 0.13; D21, 4.67 ± 0.13; *P* = 0.67), and number of secondary neurites (D20, 6 ± 0.4; D21, 6.6 ± 0.4; *P* = 0.10) present no statistical changes (Fig. 2G). When only medicated patients are included in the analysis, the number of total neurites on D21 is also higher than on D20 (D20, 9.9 ± 0.61; D21, 10.8 ± 0.61; *P* = 0.04). The rest of the structural parameter are unchanged, namely; LPN (D20, 94.1 ± 3.2µm; D21, 96.1 ± 3.2µm; *P* = 0.15), LSN (D20, 18.6 ± 0.8µm; D21, 18.9 ± 0.79µm; *P* = 0.71), number of primary neurites (D20, 4.60 ± 0.14; D21, 4.68 ± 0.14; *P* = 0.85), and number of secondary neurites (D20, 5.9 ± 0.48; D21, 6.5 ± 0.47; *P* = 0.14) (Fig. 2G).

In summary, these comparisons indicate that patients with SCZ differentiate more MDNCs (Fig. 2C) and that these MDNCs behave differently than controls cells they do not decrease its expression of CD14 (Fig. 2D) but do express high levels of nestin (Fig. 2E). In addition, MDNCs from SCZ extend longer secondary neurites and grow more primary neurites (Fig. 2F). Finally, MDNCs from patients continue to extend neurites from day 20 to 21 of the transdifferentiation process (Fig. 2G).

### Structural responses to colchicine and dopamine in MDNCs from CTL versus SCZ

We select colchicine because we have previously shown that colchicine prunes neuronal extensions from MDNCs^47^ comparably to neurons^54^ and neuronal cell lines.^55^ Moreover, colchicine acts independently of any membrane receptors and instead elicits microtubule depolymerization directly.^53^ Likewise, dopamine prunes neuronal processes in neurons during early development^43, 44, 56^ and we have found a similar response in MDNCs.^47^

MDNCs from CTL and SCZ were cultured in parallel for one hour with three concentrations of colchicine, two concentrations of dopamine and under controls conditions. For control conditions, all subjects had one sample except for one who had two samples. Correlations between microphotographs within each subject was accounted for by random effect in the mixed models. MDNCs cultured under control conditions from all groups slightly retract their LPN (CTL, 5.8 ± 1.8%; SCZ, 6.4 ± 1.6%; *P* = 0.81; MED, 5.3 ± 1.7%; *P* = 0.82) whereas, LSN grows (controls, −21.8 ± 4.4%; SCZ, −23.8 ± 3.6%; *P* = 0.73; MED, −25.6 ± 4.1%; *P* = 0.55) and neither LPN nor LSN show any differences between groups (Fig. 3A). Such culture conditions also elicit pruning of a small number of primary (CTL, 0.25 ± 0.075; SCZ, 0.27 ± 0.063; *P* = 0.47; MED, 0.24 ± 0.076; *P* = 0.57), secondary (CTL, 0.51 ± 0.16; SCZ, 0.8 ± 0.13; *P* = 0.19; MED, 0.66 ± 0.13; *P* = 0.47) and total number of neurites (CTL, 0.77 ± 0.21; SCZ, 1.22 ± 0.17; *P* = 0.12; MED, 1.04 ± 0.19; *P* = 0.35) independently of diagnosis or medications (Fig. 3A).

**Figure 3.**
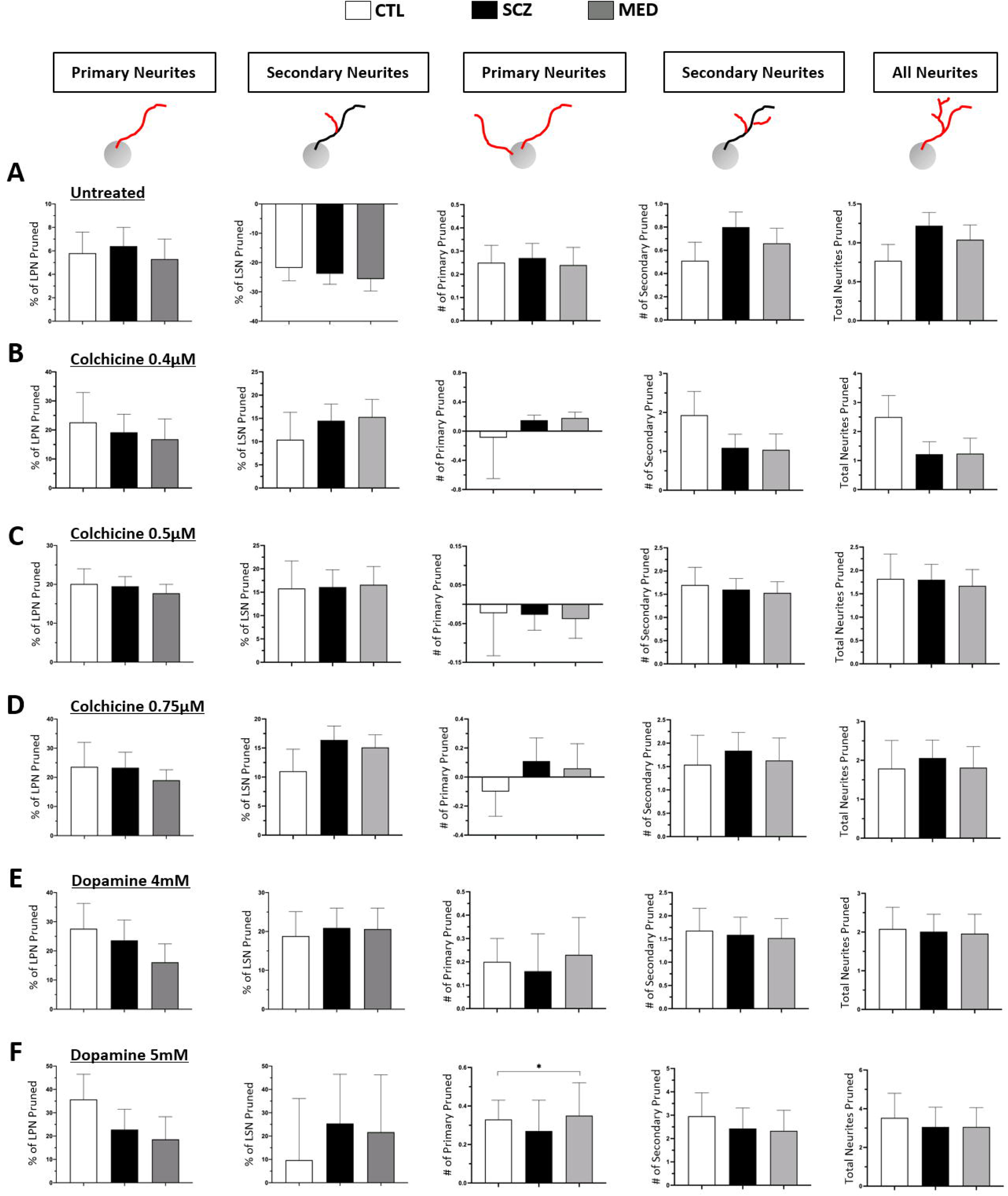
Structural responses to colchicine and dopamine in MDNCs from CTL versus SCZ. **(A)** Bar graphs contrasting the amount of pruning evidenced in MDNCs after an hour of culture under control conditions in cells from CTL, SCZ and MED. Structural parameters studied were; longest primary neurite (LPN), longest secondary neurite (LSN), number of primary neurites, number of secondary neurites and total number of neurites. Two-sample t-tests was used to determine differences between CTL vs. SCZ and CTL vs. MED after a mixed model analysis was performed to account for correlations of repeated measures within subjects. Data are given as mean ± SEM. MDNCs from 8 CTL, 10 SCZ and 8 MED were included in the analysis. MDNCs traced for LSN for CTL, *n* = 571; SCZ, *n* = 854; and MED, *n* = 667; for all other structural parameters; CTL, *n* = 656; SCZ, *n* = 976; and MED, *n* = 769. **(B)** Bar graphs contrasting the amount of pruning evidenced in MDNCs after an hour of incubation with colchicine 0.4µM in cells from CTL, SCZ and MED. The same structural parameters as in (A) were studied. We performed linear regression analysis adjusting for baseline retraction (response under control conditions), differentiation efficiency, and structure at baseline in the models as covariates based on subject-level averaged data. Data are given as mean ± SEM. MDNCs from 3 CTL, 7 SCZ and 6 MED were included in the analysis. MDNCs traced for LSN for CTL, *n* = 117; SCZ, *n* = 341; and MED, *n* = 327; for all other structural parameters; CTL, *n* = 191; SCZ, *n* = 405; and MED, *n* = 388. **(C)** Bar graphs contrasting the amount of pruning evidenced in MDNCs after an hour of incubation with colchicine 0.5µM in cells from CTL, SCZ and MED. The same structural parameters and the same statistical analysis as in (A) were used. Data are given as mean ± SEM. MDNCs from 4 CTL, 9 SCZ and 8 MED were included in the analysis. MDNCs traced for LSN for CTL, *n* = 267; SCZ, *n* = 565; and MED, *n* = 534; for all other structural parameters; CTL, *n* = 401; SCZ, *n* = 662; and MED, *n* = 627. **(D)** Bar graphs contrasting the amount of pruning evidenced in MDNCs after an hour of incubation with colchicine 0.75µM in cells from CTL, SCZ and MED. The same structural parameters and the same statistical analysis as in (A) were used. Data are given as mean ± SEM. MDNCs from 3 CTL, 7 SCZ and 6 MED were included in the analysis. MDNCs traced for LSN for CTL, *n* = 221; SCZ, *n* = 490; and MED, *n* = 472; for all other structural parameters; CTL, *n* = 297; SCZ, *n* = 621; and MED, *n* = 593. **(E)** Bar graphs contrasting the amount of pruning evidenced in MDNCs after an hour of incubation with dopamine 4mM in cells from CTL, SCZ and MED. The same structural parameters and the same statistical analysis as in (A) were used. Data are given as mean ± SEM. MDNCs from 6 CTL, 9 SCZ and 7 MED were included in the analysis. MDNCs traced for LSN for CTL, *n* = 354; SCZ, *n* = 455; and MED, *n* = 414; for all other structural parameters; CTL, *n* = 477; SCZ, *n* = 622; and MED, *n* = 486. **(F)** Bar graphs contrasting the amount of pruning evidenced in MDNCs after an hour of incubation with dopamine 5mM in cells from CTL, SCZ and MED. The same structural parameters and the same statistical analysis as in (A) were used. Data are given as mean ± SEM. MDNCs from 5 CTL, 7 SCZ and 6 MED were included in the analysis. MDNCs traced for LSN for CTL, *n* = 228; SCZ, *n* = 316; and MED, *n* = 291; for all other structural parameters; CTL, *n* = 344; SCZ, *n* = 388; and MED, *n* = 352. \**P* = or < 0.05.

To compare pruning effects of colchicine and dopamine between groups, we adjusted for baseline retraction (response under control conditions), differentiation efficiency, and structure at baseline since these factors may affect the pruning effects of each treatment condition. After accounting for these variables, we encounter that treatment with colchicine 0.4µM retracts LPN (CTL, 22.6 ± 10.3%; SCZ, 19.2 ± 6.2%; *P* = 0.80; MED, 16.8 ± 7%; *P* = 0.69) and LSN (CTL, 10.4 ± 5.9%; SCZ, 14.5 ± 3.6%; *P* = 0.60; MED, 15.3 ± 3.8%; *P* = 0.51) in all groups (Fig. 3B). In contrast, the number of primary neurites is practically unchanged (CTL, −0.089 ± 0.56; SCZ, 0.15 ± 0.07; *P* = 0.27; MED, 0.18 ± 0.08; *P* = 0.10).

Secondary neurites (CTL, 1.93 ± 0.61; SCZ, 1.09 ± 0.35; *P* = 0.33; MED, 1.04 ± 0.41; *P* = 0.33) and total number of neurites follow a similar pattern of pruning in all cohorts (CTL, 2.5 ± 0.74; SCZ, 1.2 ± 0.43; *P* = 0.23; MED, 1.24 ± 0.53; *P* = 0.30). Exposure to colchicine 0.5µM generates results comparable to those observed with colchicine 0.4µM (Fig. 3C). Colchicine at this intermediate concentration retracts LPN (CTL, 20.1 ± 3.9%; SCZ, 19.5 ± 2.5%; *P* = 0.90; MED, 17.7 ± 2.3%; *P* = 0.67) and LSN (CTL, 15.8 ± 5.9%; SCZ, 16.1 ± 3.7%; *P* = 0.97; MED, 16.6 ± 3.9%; *P* = 0.89) equally in every cohort. In close resemblance to colchicine 0.4µM, colchicine 0.5µM elicits minimal changes in primary neurites (CTL, −0.023 ± 0.11; SCZ, −0.027 ± 0.04; *P* = 0.59; MED, −0.038 ± 0.05; *P* = 0.33), while the three groups retract a similar number of secondary neurites (CTL, 1.7 ± 0.38; SCZ, 1.6 ± 0.24; *P* = 0.24; MED, 1.53 ± 0.24; *P* = 0.63) and total number of neurites (CTL, 1.82 ± 0.53; SCZ, 1.80 ± 0.33; *P* = 0.97; MED, 1.67 ± 0.35; *P* = 0.80). Results from the highest concentration of colchicine tested (0.75µM) follow a similar pattern (Fig. 3D). Colchicine 0.75µM reduces the size of LPN (CTL, 23.6 ± 8.4%; SCZ, 23.3 ± 5.3%; *P* = 0.97; MED, 19.0 ± 3.6%; *P* = 0.58) and LSN (CTL, 11 ± 3.8%; SCZ, 16.4 ± 2.4%; *P* = 0.30; MED, 15.1 ± 2.2%; *P* = 0.37) comparably in all cohorts, while the number of primary neurites remains stable (CTL, −0.1 ± 0.17; SCZ, 0.11 ± 0.16; *P* = 0.71; MED, 0.06 ± 0.17; *P* = 0.81). Secondary neurites (CTL, 1.5 ± 0.63; SCZ, 1.8 ± 0.39; *P* = 0.72; MED, 1.63 ± 0.48; *P* = 0.83) and total number of neurites (CTL, 1.7 ± 0.72; SCZ, 2 ± 0.46; *P* = 0.77; MED, 1.81 ± 0.54; *P* = 0.83) retract at equivalent rates independently of diagnosis or medications.

MDNCs from the three groups incubated with dopamine 4mM retract its LPN (CTL, 27.6 ± 8.7%; SCZ, 23.6 ± 7%; *P* = 0.73; MED, 16.1 ± 6.3%; *P* = 0.23) and LSN (CTL, 18.8 ± 6.3%; SCZ, 20.9 ± 5.1%; *P* = 0.81; MED, 20.6 ± 5.4%; *P* = 0.93) comparably (Fig. 3E). Likewise, pruning of primary (CTL, 0.20 ± 0.10; SCZ, 0.16 ± 0.16; *P* = 0.51; MED, 0.23 ± 0.16; *P* = 0.51), secondary (CTL, 1.6 ± 0.48; SCZ, 1.59 ± 0.38; *P* = 0.89; MED, 1.52 ± 0.42; *P* = 0.72) and total number of neurites (CTL, 2.08 ± 0.56; SCZ, 2.01 ± 0.45; *P* = 0.93; MED, 1.96 ± 0.5; *P* = 0.71) reveals no statistical differences. There are also no statistical differences in the reduction of LPN (CTL, 35.7 ± 10.8%; SCZ, 22.8 ± 8.7%; *P* = 0.43; MED, 18.6 ± 9.7%; *P* = 0.31) or LSN (CTL, 9.7 ± 26.4%; SCZ, 25.4 ± 21.1%; *P* = 0.69; MED, 21.7 ± 24.6%; *P* = 0.78) after treatment with dopamine 5mM (Fig. 3F). In contrast, this higher concentration of dopamine prunes a slightly higher but statistically significant number of primary neurites in MDNCs from MED but not from SCZ when compared with CTL (CTL, 0.33 ± 0.10; SCZ, 0.27 ± 0.16; *P* = 0.52; MED, 0.35 ± 0.17; *P* = 0.05) (Fig. 3F). No significant differences are evident in the loss of secondary (CTL, 2.96 ± 1.0; SCZ, 2.43 ± 0.88; *P* = 0.74; MED, 2.33 ± 0.88; *P* = 0.62) or total number of neurites (CTL, 3.53 ± 1.27; SCZ, 3.06 ± 1.02; *P* = 0.80; MED, 3.06 ± 1.0; *P* = 0.72).

In summary, MDNCs from medicated patients with SCZ prune more primary neurites when treated with dopamine 5mM than cells from controls (Fig. 3F). There were no other differences in the structural response of MDNCs after exposure to a lower concentration of dopamine and after treatment with three different concentrations of colchicine (Fig. 3).

### Dopamine 1 receptors in MDNCs from controls versus SCZ

Activation of D1R has been consistently associated with pruning of neuronal extensions during early stages of development *in vivo*^41^ and *in vitro*.^43^ We have previously measured expression of D1R in MDNCs from healthy individuals.^47^ Here we compare expression of D1R in MDNCs from CTL versus SCZ. But first, we replicated previous reports indicating that monocytes do not express D1R^57^ (Fig. 4A). In contrast, MDNCs from CTL and SCZ express D1R suggesting that our transdifferentiation protocol promotes its expression (Fig. 4B). Expression of D1R in MDNCs is not statistically different between CTL and patients with SCZ (CTL, 46.4 ± 6.7; SCZ, 31.3 ± 5.2; *P* = 0.15) (Fig. 4C). However, if only medicated patients are included in the analysis, the difference becomes significant with patients expressing less D1R (CTL, 46.4 ± 6.7; MED, 24.2 ± 3.4; *P* = 0.03) (Fig. 4C).

**Figure 4.**
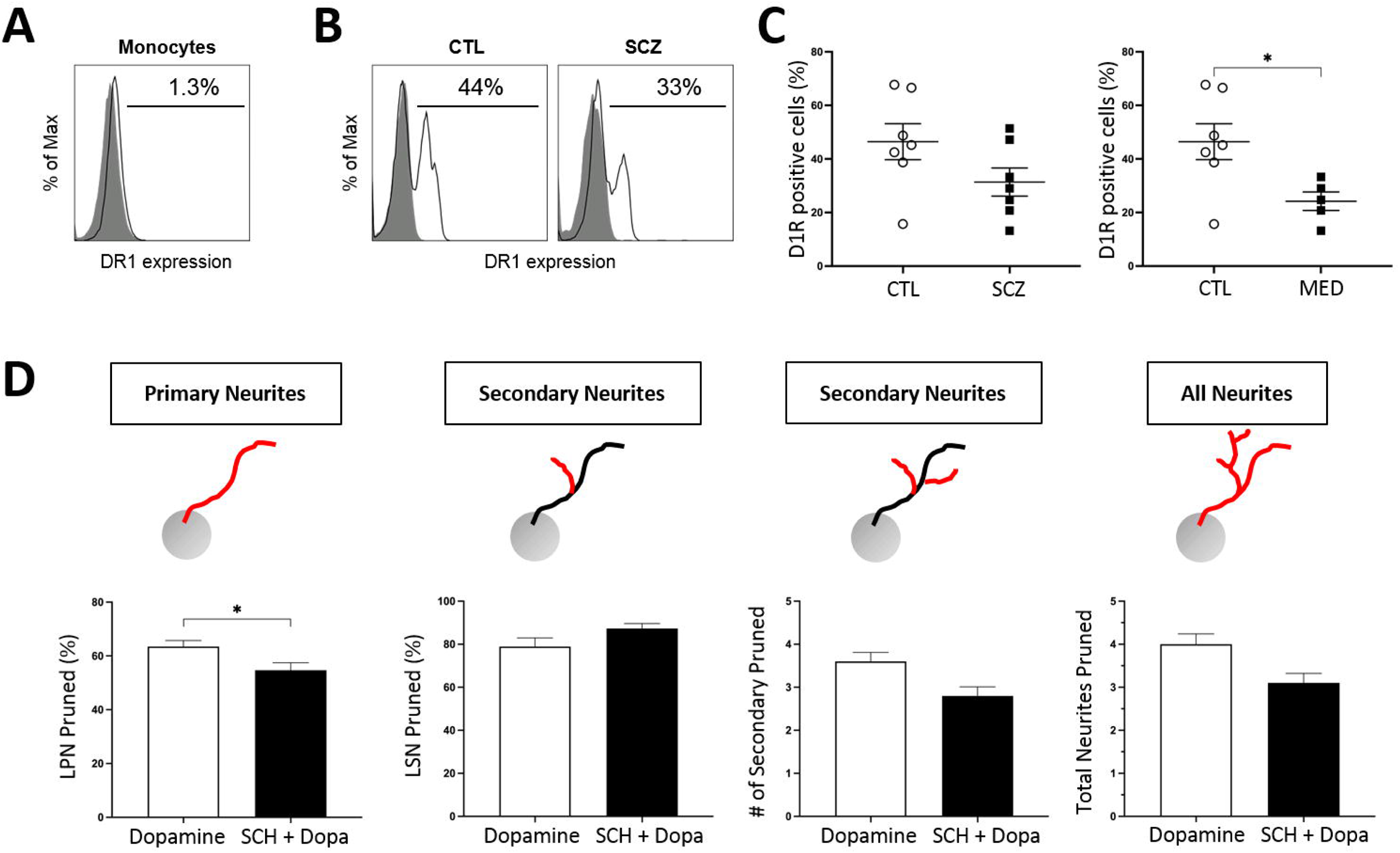
Dopamine 1 receptors in MDNCs from CTL and SCZ. **(A)** Flow cytometric diagram showing expression of dopamine 1 receptors (D1R) in monocytes. Gray histograms represent control isotypic labeling. White histograms represent specific labeling. **(B)** Flow cytometric diagrams showing expression of D1R in MDNCs from a healthy control and a patient with SCZ. Gray histograms represent control isotypic labeling. White histograms represent specific labeling. **(C)** Dot plots contrasting expression of D1R in MDNCs from CTL vs. SCZ and CTL vs. MED. Non-parametric Mann-Whitney test was used to make pairwise comparisons between groups. Data are given as mean ± SEM. Cells from 7 CTL, 7 SCZ and 5 MED were included in the analysis. \**P* = or < 0.05. **(D)** Bar graphs showing the effects of SCH-23390, a D1R antagonist, in dopamine-elicited pruning in MDNCs. The non-parametric Mann-Whitney test was used to make pairwise comparisons between groups. Data are given as mean ± SEM. MDNCs traced for LSN treated with dopamine *n* = 200; and with SCH-23390 + dopamine, *n* = 117; for all other structural parameters; treated with dopamine *n* = 221; and with SCH-23390 + dopamine, *n* = 125. \**P* = or < 0.05.

We then tested whether preincubation with SCH-23390, a D1R antagonist, prevents any of the structural changes elicited by dopamine (Fig. 4D). SCH-23390 decreases pruning in LPN by 10% which is statistically significant (dopamine, 63.5 ± 2.2%; SCH, 54.7 ± 2.8%; *P* = 0.0093). In contrast, pruning of LSN is not significantly affected by SCH-23390 (dopamine, 78.9 ± 4.0%; SCH, 87.3 ± 2.3%; *P* = 0.93). No primary neurites were pruned either with dopamine or SCH-23390 + dopamine, while the number of secondary neurites (dopamine, 3.6 ± 0.21; SCH, 2.8 ± 0.21; *P* = 0.08) and total number of neurites (dopamine, 4.0 ± 0.24; SCH, 3.1 ± 0.22; *P* = 0.08) tends to decrease in the presence of SCH-23390 but without reaching statistical significance (Fig. 4D).

### Haloperidol effects on the structure and D1R expression of MDNCs

In order to determine whether antipsychotics impact levels of differentiation or the structure of MDNCs, we incubated monocytes from day 4 to 7 of the transdifferentiation process with haloperidol similar to known circulating levels^58^ (20ng/ml). Haloperidol was selected because three patients with the highest differentiation efficiency were taking this antipsychotic. The incubation period consisting of 3 days was chosen to resemble in vivo conditions, where human monocytes remain in circulation for around 2 days.^59^

A non-parametric one-way ANOVA on the percentage of differentiated MDNCs established by phenotype, indicates CTL cells are not statistically different to those incubated with either vehicle (VEH) or haloperidol (HAL) (CTL, 6.2 ± 0.58%; VEH, 7.5 ± 0.93%; HAL, 6.8 ± 0.65%; *P* = 0.69) (Fig. 5A). The structure of MDNCs also remains unchanged regardless of treatment (Fig. 5B). Our structural analysis includes; LPN (CTL, 68.7 ± 1.5µm; VEH, 65.8 ± 1.8µm; HAL, 66 ± 2µm; *P* = 0.52), LSN (CTL, 14.6 ± 0.9µm; VEH, 13.8 ± 0.8µm; HAL, 13.4 ± 0.5µm; *P* = 0.65), number of primary (CTL, 3.1 ± 0.1; VEH, 3.2 ± 0.07; HAL, 2.9 ± 0.09; *P* = 0.08), secondary (CTL, 2.1 ± 0.1; VEH, 2.1 ± 0.1; HAL, 1.9 ± 0.1; *P* = 0.59), and total number of neurites (CTL, 5.5 ± 0.2; VEH, 5.4 ± 0.2; HAL, 4.9 ± 0.2; *P* = 0.27) (Fig. 5B).

**Figure 5.**
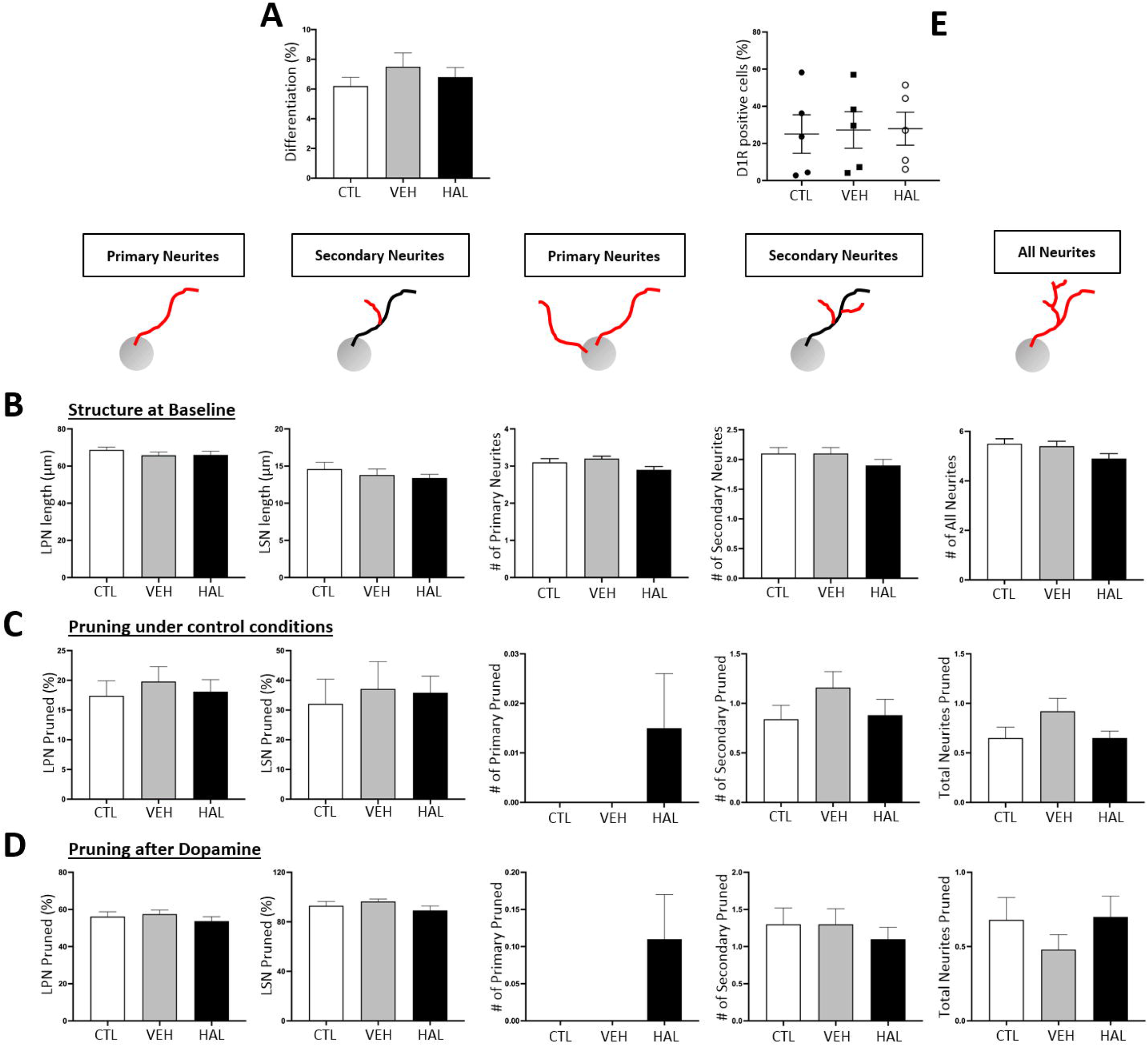
Haloperidol effects on the structure and D1R expression of MDNCs. **(A)** Bar graphs contrasting the differentiation percentage between cells treated with haloperidol (HAL), vehicle (VEH) or under control conditions (CTL). The Kruskal-Wallis Test was used to make comparisons between groups. Data are given as mean ± SEM. Cells from 5 healthy subjects were included in the analysis. Cells characterized for CTL, *n* = 8048; VEH, *n* = 7152; HAL, *n* = 9300. **(B)** Bar graphs contrasting the structure of MDNCs at baseline after treatment from day 4 to 7 of the transdifferentiation process with haloperidol, vehicle or cells under control conditions. Structural parameters studied were: longest primary neurite (LPN), longest secondary neurite (LSN), number of primary neurites, number of secondary neurites and total number of neurites. The Kruskal-Wallis Test was used to make comparisons between groups. Data are given as mean ± SEM. Cells from 4 healthy subjects were included in the analysis. MDNCs traced for LSN for CTL, *n* = 247; VEH, *n* = 278; HAL, *n* = 305; for all other structural parameters; CTL, *n* = 339; VEH, *n* = 382; HAL, *n* = 431. **(C)** Bar graphs contrasting the amount of pruning evidenced in MDNCs after an hour of incubation on cells treated from day 4 to 7 of the transdifferentiation process with haloperidol, vehicle or cells under control conditions. The same structural parameter as in (B) were assessed. The Kruskal-Wallis Test was used to make comparisons between groups. Data are given as mean ± SEM. Cells from 3 healthy subjects were included in the analysis. MDNCs traced for LSN for CTL, *n* = 64; VEH, *n* = 85; HAL, *n* = 124; for all other structural parameters; CTL, *n* = 98; VEH, *n* = 113; HAL, *n* = 189. **(D)** Bar graphs contrasting the amount of pruning evidenced in MDNCs after an hour of incubation with dopamine 5mM in cells treated from day 4 to 7 of the transdifferentiation process with haloperidol, vehicle or cells under control conditions. The same structural parameter as in (B) were assessed. The Kruskal-Wallis Test was used to make comparisons between groups. Data are given as mean ± SEM. Cells from 2 healthy subjects were included in the analysis. MDNCs traced for LSN for CTL, *n* = 28; VEH, *n* = 23; HAL, *n* = 37; for all other structural parameters; CTL, *n* = 57; VEH, *n* = 68; HAL, *n* = 81. **(E)** Dot plots contrasting expression of D1R in MDNCs after treatment from day 4 to 7 of the transdifferentiation process with haloperidol, vehicle or cells under control conditions. The Kruskal-Wallis Test was used to make comparisons between groups. Data are given as mean ± SEM. Cells from 5 healthy subjects were included in the analysis.

We also studied whether haloperidol treatment impacts pruning (Fig. 5C). LPN from MDNCs cultured under CTL conditions, VEH or HAL retract equally (CTL, 17.4 ± 2.5%; VEH, 19.8 ± 2.5%; HAL, 18.1 ± 2%; *P* = 0.92). Likewise, the amount of retraction in LSN between the three culture conditions is similar (CTL, 32.1 ± 8.3%; VEH, 37.1 ± 9.2%; HAL, 35.9 ± 5.5%; *P* = 0.48). The number of primary (CTL, 0 ± 0; VEH, 0 ± 0; HAL, 0.015 ± 0.011; *P* = 0.98), secondary neurites (CTL, 0.84 ± 0.14; VEH, 1.16 ± 0.16; HAL, 0.88 ± 0.16; *P* = 0.33) and total number of pruned neurites (CTL, 0.65 ± 0.11; VEH, 0.92 ± 0.13; HAL, 0.65 ± 0.07; *P* = 0.24) is comparable between the three groups (Fig. 5C).

The pruning effects of dopamine 5mM on MDNCs was also investigated after haloperidol treatment from day 4 to 7 (Fig. 5D). Pruning of LPN (CTL, 56.2 ± 2.5%; VEH, 57.5 ± 2.2%; HAL, 53.7 ± 2.4%; *P* = 0.54) and LSN (CTL, 93 ± 3.4%; VEH, 96.4 ± 2%; HAL, 89.2 ± 3.6%; *P* = 0.75) is equivalent between the three treatment conditions. The same is true for number of primary (CTL, 0 ± 0; VEH, 0 ± 0; HAL, 0.015 ± 0.011; *P* = 0.83), secondary (CTL, 1.3 ± 0.22; VEH, 1.3 ± 0.21; HAL, 1.1 ± 0.16; *P* = 0.79) and total number of pruned neurites (CTL, 0.68 ± 0.15; VEH, 0.48 ± 0.1; HAL, 0.7 ± 0.14; *P* = 0.54) (Fig. 5D). Finally, we measured whether haloperidol alters the expression of D1R. MCNCs’ expression of D1R is unaffected by HAL treatment (CTL, 25 ± 10.3%; VEH, 27.2 ± 9.8%, HAL, 27.9 ± 8.8%; P = 0.87) (Fig. 5E).

In summary, incubation of haloperidol from day 4 to 7 of the transdifferentiation process does not alter the neurostructure of MDNCs or influences pruning elicited by dopamine. Expression of D1R by MDNCs is also unaffected by haloperidol exposure from day 4 to 7.

## Discussion

Despite intense research during the last several decades, the pathophysiology of SCZ remains obscure. One of the main obstacles to study this mental illness is the difficulty of accessing neurons directly from patients. The extended lapse between SCZ putative origin during neurodevelopment and its clinical emergence makes this challenge even harder. In order to overcome this obstacle, we used MDNCs that allow us to test *in vitro* whether some of the very early neurodevelopmental steps involving the neuronal structure are deficient in SCZ. Particularly intriguing is the possibility that patients with SCZ carry an increased susceptibility to the pruning effects of dopamine. This susceptibility is likely to be more obvious in the early stages of neuronal development when dopamine pruning effects appear to be more prominent.^40–46^ In fact, dopamine removes neuronal extensions even before commitment into any particular neuronal type is yet achieved.^40, 42, 44, 45^ MDNCs stand within that neurodevelopmental window.^47^ Also significant is that MDNCs replicate other characteristics found in HDNs.

In a prior publication we showed that the structure of MDNCs is comparable to that of HDNs maintained in culture for 5 days.^47^ As a further step to characterize the neurostructure of MDNCs, here we contrasted how MDNCs and HDNs respond to colchicine, a compound well-known in neuroscience for its ability to prune neuronal extensions.^52^ MDNCs reproduce changes in LPN, number of primary neurites and total number of neurites found in HDNs (Fig. 1C). In contrast, MDNCs do not replicate variations in LSN or number of secondary neurites. These results suggest that findings from MDNCs involving pruning of LPN, number of primary neurites and/or total number of neurites are likely to be found in HDNs, while insights relating to secondary neurites would not.

Concerns have been raised about the reproducibility of results obtained with other cellular models.^48, 49, 60^ Given these concerns, we have previously reported that MDNCs deliver reproducible results in a small cohort of healthy men.^47^ Here we expanded our cohort to include women and we also increased the number of samples tested (Table 2). In our current cohort, reproducibility on the percentage of differentiated MDNCs is low, while the number of differentiated cells is consistent (Fig. 1A). A plausible explanation for these results is based on previous reports indicating that the capacity to transdifferentiate is limited to a specific type of monocyte.^61^ A ceiling effect may at play that limits the production of such monocytes.

We also tested the reproducibility of several neurostructural parameters and found different degrees of consistency (Fig. 1A). Results involving LPN were more consistent that those relating to LSN, number of secondary neurites and total number of neurites. Reproducibility for number of primary neurites was low, found only in 60% of the subjects tested. Taking together the neurostructural comparison between MDNCs and HDNs (Fig. 1C) as well as the reproducibility of MDNCs (Fig. 1A), we conclude that differences between patients and controls that pertain to LPN and total number of neurites should be considered reliable. Results involving any of the other structural parameters studied, should be deemed less reliable, as they are either not found in HDNs or not consistently encountered in MDNCs.

The cohort of patients with SCZ and controls we recruited was matched by age and gender (Table 1). The number of PBMCs and monocytes did not differ between cohorts. However, patients presented a small but statistically significant increase in the percentage of monocytes within PBMCs (Table 3). These results are consistent with several reports indicating patients with SCZ have higher numbers of monocytes.^62^ The reason for this monocytosis might be related to some degree of inflammation present in patients with schizophrenia, as different publications have associated this psychotic disorder with plasmatic, transcriptomic and epigenetic signs of inflammation,^63^ including complement, coagulation factors,^64^ and lipidomic changes^65^ but this possibility remains to be established.

While the path to differentiation is similar between groups (Fig. 2B), MDNCs from patients with SCZ differentiate more efficiently (Fig. 2C) and develop a more elaborated structure, presenting a higher number of primary neurites and longer secondary neurites (Fig. 2F). Previous publications using Induced Pluripotent Stem Cells (IPSCs) from patients with SCZ have also reported a more complex structure during early stages of neurodevelopment^66^ though, not always.^26^

In a previous publication we showed via genetic, immunofluorescence and flow cytometry that MDNCs decrease its expression of CD14 when compared to macrophages from the same individual.^47^ A drop that was expected since CD14 is a marker for monocytes/macrophages. However, MDNCs from patients did not behave as anticipated. We observed a small decrease in the expression of CD14 that was not significant (Fig. 2D). In contrast, MDNCs from CTL present a pronounced and statistically significant reduction in this marker for monocytes/macrophages (Fig. 2D). The structure of MDNCs from patients also behaves differently. While the neurostructure of MDNCs from CTL remains stable after the transdifferentiation process is completed, cells from patients continue to grow neurites during the following 24 hours (Fig. 2G).

After accounting in our statistical analysis for differences between SCZ and CTL in differentiation efficiency, size of secondary neurites and number of primary neurites, we challenged the structure of MDNCs’ with colchicine and dopamine. Both compounds are capable of pruning neuronal extensions through different mechanisms of action. Colchicine acts by directly depolymerizing microtubules^53^ and thus, it is independent of membrane receptors, whereas dopamine relies on activation of D1R.^41, 43^ Three different concentrations of colchicine known to elicit pruning of extensions in neurons^54^ and neuronal cell lines,^55^ did not reveal any changes between MDNCs from SCZ and CTL (Fig. B-D). These results indicate that the microtubular component of the cytoskeleton is unaffected in MDNCs from patients with SCZ. Our data contrasts with previous reports suggesting patients with SCZ present deficits in microtubules organization and stability, granted those experiments were conducted using olfactory neuroepithelial cells.^67, 68^

Two different concentrations of dopamine did not reveal differences in LPN, LSN, number of secondary neurites or total number of neurites between groups (Fig. E, F). Nonetheless, our experiments using SCH-23390, a specific D1R antagonist, indicate that at the concentrations of dopamine we tested, this receptor is involved in pruning of LPN (Fig. 4C). Expression of D1R in MDNCs was similar in SCZ and CTL (Fig. 4B). When only medicated patients are included in the analysis, expression of D1R is significantly lower in patients with SCZ (Fig. 4B). It is therefore possible, that in medicated patients an increased susceptibility to the pruning effects of dopamine in LPN was masked by lower levels of D1R. MDNCs from medicated patients also evidenced increased pruning of primary neurites after exposure to the higher dose of dopamine (Fig. 3F). These results suggest that MDNCs from medicated patients are more susceptible to pruning of primary neurites and plausibly also to a partial removal of LPN by dopamine. It has to be kept in mind however, that the reliability of data involving primary neurites from MDNCs is low because of the variability of results with serial samples. In addition, the potential contribution of antipsychotics in the pruning processes appears likely. But several factors need to be pondered before drawing any conclusions.

Monocytes are exposed to medications during the 2-3 days they remain in blood circulation.^59^ After monocyte extraction and while in culture for 20 days, there is no further exposure to medications. In fact, cell culture media is replaced in 4 occasions during the transdifferentiation process removing monocytes even further from their circulating environment.^47^ Another important aspect to consider is that monocytes do not express dopamine receptors.^57^ Here we replicated the absence of D1R expression in monocytes (Fig. 4A). Consequently, any antipsychotic influence would have to take place independently of dopamine receptors. Furthermore, incubations with haloperidol that mimic *in vivo* exposure, did not impact MDNCs’ differentiation, neurostructure or D1R expression (Fig. 5A-E). It is nonetheless possible, that antipsychotics could exert its effects via epigenetic mechanisms.^69, 70^ Another potential alternative is that our findings involving only medicated patients could be due to patient heterogeneity rather than to medication effect. Further studies are needed to confirm this statement. In the meantime, the effects of medications on any of our comparisons between SCZ and CTL should be considered a confounding factor.

There is additional information that needs to be highlighted to properly appraise our results. While our cohort is among the largest in the field of SCZ and stem cells, only a subset of individuals were included when we tested MDNCs structural responses to colchicine and dopamine. MDNCs from 3 to 10 individuals from each group were included in these analyses (Fig. 3). Thus, the cohorts for some of these subset analyses are small. It is also important to consider that we used supra-physiological concentrations of dopamine to elicit rapid pruning of neuronal extensions. Consequently, our *in vitro* culture conditions do not resemble the microenvironment that surrounds neurons *in vivo*. Nonetheless, *in vitro* experiments using cells that carry the genetic susceptibility to SCZ and deliver results in only 20 days, open the possibility to a future cellular-based characterization of patients which is greatly needed in the psychiatric field.

## Author contributions

AB planned and developed most experiments and wrote this manuscript, VF participated in the planning, development and analysis of several experiments, ACR developed and helped planned some of the experiments, FM participated in the development of the discussion, LK developed most of the statistical analysis and participated in the interpretation of the results, LEH participated in the interpretation of the results and the development of the discussion, LDG developed the analysis of some experiments, TMJ participated in the planning of several experiments, AH participated in the planning and interpretation of experiments and in the writing of this manuscript, MOK participated in the planning of experiments, participant selection and in the development of this manuscript.

## Data Availability

All data produced in the present study are available upon reasonable request to the authors.

## Acknowledgments

We would like to thank Professor Andrew Francis for his thorough editorial comments. We also would like to thank the Clinical Research Team from Sainte-Anne Hospital (CERC) for their technical assistance as well as Xinyi Liu for her input on statistics. This work was financially supported by Penn State College of Medicine, Penn State Hershey Medical Center, Université Paris Descartes Sorbonne Paris Cité, Institut National de la Sante et de la Recherche Medicale (INSERM), CNRS, Foundation pour la Recherche Medicale (FRM)(SPF20080511940 to AB; FRM DPP 2015 1033966 to MOK, AH and AW), the French Government’s Investissement d’Avenir program, Laboratoires d’Excellence “Integrative Biology of Emerging Infectious Diseases” (ANR-10-LABX-62-IBEID), Consejo Nacional de Ciencia y Tecnologia (CONACYT 74641 to AB), by the Young Minds in Psychiatry Award given by the American Psychiatric Association to AB and by the Ling and Esther Tan Early Career Professor Award given to AB by the Department of Psychiatry and Behavioral Health, Penn State Hershey Medical Center.

## Figure Legends

**Supplemental Table 1.** Reproducibility of MDNCs’ differentiation percentage after serial samples from healthy individuals.

| Differentiation Percentage |  |  |  |
| --- | --- | --- | --- |
| Subject | Difference Mean | 95% CL range | Satterthwaite test |
| S1 | -0.031 | -0.061 to -0.0009 | $P = 0.04$ |
| S2 | 0.040 | 0.015 to 0.065 | $P = 0.003$ |
| S3 | -0.090 | -0.145 to -0.036 | $P = 0.002$ |
| S4 | -0.014 | -0.047 to 0.020 | $P = 0.38$ |
| S5 | -0.047 | -0.102 to 0.008 | $P = 0.08$ |
| S6 | -0.042 | -0.072 to -0.012 | $P = 0.007$ |
| S7 | 0.012 | -0.042 to 0.066 | $P = 0.63$ |
| S8 | 0.013 | -0.001 to 0.027 | $P = 0.07$ |

CL = confidence interval

**Supplemental Table 2.** Reproducibility in number of differentiated MDNCs after serial samples from healthy individuals.

| Number of differentiated MDNCs |  |  |  |
| --- | --- | --- | --- |
| Subject | Difference Mean | 95% CL range | Satterthwaite test |
| S1 | 0.312 | -0.508 to 1.132 | $P = 0.43$ |
| S2 | 0.461 | -0.054 to 0.976 | $P = 0.07$ |
| S3 | 0.244 | -0.264 to 0.752 | $P = 0.32$ |
| S4 | 0.333 | -0.946 to 1.613 | $P = 0.58$ |
| S5 | 0.577 | -0.418 to 1.572 | $P = 0.23$ |
| S6 | 0.741 | 0.220 to 1.262 | $P = 0.006$ |
| S7 | 0.566 | -0.525 to 1.656 | $P = 0.28$ |
| S8 | -0.351 | -0.899 to 0.196 | $P = 0.20$ |

CL = confidence interval

**Supplemental Table 3.** Reproducibility in longest primary neurite (LPN) from MDNCs after serial samples from healthy individuals.

| Longest Primary Neurite |  |  |  |
| --- | --- | --- | --- |
| Subject | Difference Mean | 95% CL range | Satterthwaite test |
| S1 | 0.33 | -17.36 to 16.69 | $P = 0.96$ |
| S2 | 1.94 | -23.97 to 27.85 | $P = 0.87$ |
| S3 | 5.4 | -21.48 to 32.3 | $P = 0.68$ |
| S4 | 1.79 | -14.58 to 18.17 | $P = 0.82$ |
| S5 | -9.27 | -25.27 to 6.72 | $P = 0.24$ |
| S6 | -3.88 | -17.54 to 9.77 | $P = 0.57$ |
| S7 | 2.56 | -16.53 to 21.65 | $P = 0.78$ |
| S8 | 5.01 | -7.16 to 17.19 | $P = 0.41$ |

CL = confidence interval

**Supplemental Table 4.**
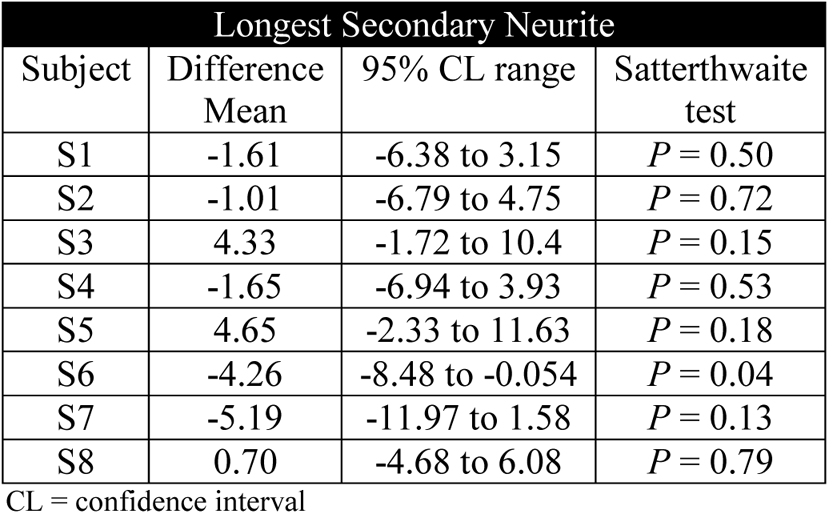
Reproducibility in longest secondary neurite (LSN) from MDNCs after serial samples from healthy individuals.

| Longest Secondary Neurite |  |  |  |
| --- | --- | --- | --- |
| Subject | Difference Mean | 95% CL range | Satterthwaite test |
| S1 | -1.61 | -6.38 to 3.15 | $P = 0.50$ |
| S2 | -1.01 | -6.79 to 4.75 | $P = 0.72$ |
| S3 | 4.33 | -1.72 to 10.4 | $P = 0.15$ |
| S4 | -1.65 | -6.94 to 3.93 | $P = 0.53$ |
| S5 | 4.65 | -2.33 to 11.63 | $P = 0.18$ |
| S6 | -4.26 | -8.48 to -0.054 | $P = 0.04$ |
| S7 | -5.19 | -11.97 to 1.58 | $P = 0.13$ |
| S8 | 0.70 | -4.68 to 6.08 | $P = 0.79$ |

CL = confidence interval

**Supplemental Table 5.** Reproducibility in number of primary neurites from MDNCs after serial samples from healthy individuals.

| Number of Primary Neurites |  |  |  |
| --- | --- | --- | --- |
| Subject | Difference Mean | 95% CL range | Satterthwaite test |
| S1 | 1.9 | 1.02 to 2.79 | $P = 0.0001$ |
| S2 | 1.82 | 0.62 to 3.02 | $P = 0.005$ |
| S3 | 0.12 | -0.9 to 1.14 | $P = 0.81$ |
| S4 | 0.29 | -0.53 to 1.12 | $P = 0.47$ |
| S5 | 0.06 | -1.16 to 1.28 | $P = 0.91$ |
| S6 | -1.07 | -1.74 to 0.4 | $P = 0.002$ |
| S7 | 0.26 | -0.88 to 1.42 | $P = 0.64$ |
| S8 | -0.34 | -1.13 to 0.44 | $P = 0.38$ |

CL = confidence interval

**Supplemental Table 6.** Reproducibility in number of secondary neurites from MDNCs after serial samples from healthy individuals.

| Number of Secondary Neurites |  |  |  |
| --- | --- | --- | --- |
| Subject | Difference Mean | 95% CL range | Satterthwaite test |
| S1 | 0.088 | -2.4 to 2.58 | $P = 0.94$ |
| S2 | -0.27 | -4.14 to 3.6 | $P = 0.88$ |
| S3 | -0.41 | -3.36 to 2.53 | $P = 0.77$ |
| S4 | -1.28 | -2.94 to 0.38 | $P = 0.12$ |
| S5 | 0.58 | -3.43 to 4.59 | $P = 0.77$ |
| S6 | -3.57 | -5.69 to 1.46 | $P = 0.001$ |
| S7 | -0.75 | -3.98 to 2.48 | $P = 0.64$ |
| S8 | -0.92 | -2.91 to 1.06 | $P = 0.35$ |

CL = confidence interval

**Supplemental Table 7.** Reproducibility in total number of neurites from MDNCs after serial samples from healthy individuals.

| Total Number of Neurites |  |  |  |
| --- | --- | --- | --- |
| Subject | Difference Mean | 95% CL range | Satterthwaite test |
| S1 | 2.51 | -0.89 to 5.91 | $P = 0.14$ |
| S2 | 1.0 | -4.42 to 6.45 | $P = 0.70$ |
| S3 | 0.54 | -3.08 to 4.16 | $P = 0.76$ |
| S4 | -1.55 | -4.19 to 1.09 | $P = 0.24$ |
| S5 | 0.77 | -4.89 to 6.44 | $P = 0.78$ |
| S6 | -5.88 | -8.91 to -2.85 | $P = 0.0002$ |
| S7 | -1.21 | -5.93 to 3.5 | $P = 0.60$ |
| S8 | -1.05 | -4.02 to 1.91 | $P = 0.48$ |

CL = confidence interval

**Supplemental Table 8.** Structural path to transdifferentiation in patients with schizophrenia and control individuals.

| Day | Structural Stage | Controls | Patients with schizophrenia | ANOVAs |
| --- | --- | --- | --- | --- |
| Day 4 | RC | 32.3 ± 4.6% | 33.3 ± 4.4% | *P* = 0.88 |
|  | SM | 10.5 ± 2.3% | 11.6 ± 2.2% | *P* = 0.72 |
|  | FS | 29.9 ± 5% | 34 ± 4.9% | *P* = 0.56 |
|  | UC | 27.1 ± 2.9% | 20.9 ± 2.8% | *P* = 0.14 |
| Day 7 | RC | 22 ± 4.8% | 13.7 ± 4.6% | *P* = 0.22 |
|  | SM | 23.2 ± 5.7% | 26.4 ± 5.5% | *P* = 0.69 |
|  | FS | 32.6 ± 5.1% | 44.2 ± 4.9% | *P* = 0.11 |
|  | UC | 21.9 ± 1.9% | 15.5 ± 1.8% | *P* = 0.02 |
| Day 10 | RC | 14.8 ± 4% | 12.2 ± 3.9% | *P* = 0.64 |
|  | SM | 36.6 ± 7.1% | 34.4 ± 6.9% | *P* = 0.82 |
|  | FS | 28.1 ± 5.5% | 35.1 ± 5.3% | *P* = 0.36 |
|  | UC | 20.3 ± 2.4% | 18.1 ± 2.4% | *P* = 0.52 |
| Day 13 | RC | 13.5 ± 3.5% | 10.4 ± 2.4% | *P* = 0.47 |
|  | SM | 29.9 ± 6.5% | 37.6 ± 4.4% | *P* = 0.19 |
|  | FS | 41 ± 7.1% | 27.9 ± 4.8% | *P* = 0.15 |
|  | UC | 18.3 ± 3.1% | 23.9 ± 2% | *P* = 0.15 |

RC = rounded cell, SM = standard macrophage, FS = fibroblastic shape and UC = uncharacterized cells.

